# Non-linear Relationships between COVID-19 and Non-COVID-19 Mortality by Vaccination Status within Age Groups

**DOI:** 10.1101/2025.08.22.25333889

**Authors:** Ir. A.J. Oostenbrink

## Abstract

**Background:** Observational studies on COVID-19 vaccine effectiveness (VE) are prone to biases such as healthy vaccinee effect and misclassification, potentially distorting mortality patterns by vaccination status. The proportionality hypothesis posits that COVID-19 mortality is proportional to all-cause mortality within subgroups, but selection bias and frailty may introduce non-linearity.

**Objective:** To investigate non-linear relationships between COVID-19 and non-COVID-19 mortality by vaccination status using UK data, modeling with a power function to correct for biases.

**Methods:** Monthly age-standardized mortality rates from the Office for National Statistics (ONS) database (January 2021–May 2023) were analyzed for five vaccination statuses and six age groups. Relative risks (RR) were calculated, and a power function (RRcov ∝ (RRnoncov)^a) was used to test non-linearity. Optimal a-values were validated via visual analysis of VE curves, focusing on stability and logical patterns.

**Results:** In older age groups (≥70 years), the relationship was non-linear (a ≈1.5–2), with VE stabilizing at 60–90% after correction. No significant improvement from additional vaccination doses was observed. Younger groups showed near-linearity (a ≈1–1.5). Concentration effects explained higher mortality in groups not receiving additional doses.

**Conclusions:** The non-linear model corrects for biases, suggesting initial vaccination suffices for protection and multiple boosters may be unnecessary. Frailty likely drives this non-linearity in older groups. However, visual analysis limits robustness; future validation with individual-level data and statistical tests is needed.

## Introduction

The analysis of mortality patterns during the COVID-19 pandemic is complicated by selection bias and dynamic changes in population compositions, particularly due to vaccination status. Studies have shown that biases such as the healthy vaccinee bias and potential misclassification of vaccination status lead to apparent differences in mortality between vaccinated and unvaccinated groups (Bakker et al., 2025; Neil et al., 2022). These biases arise in part from self-selection, where terminally ill individuals (hereafter “rapidly dying”) or those recently infected with COVID-19 do not get vaccinated, resulting in a concentration of vulnerable individuals in the non-(further)-vaccinated groups.

Frailty, as a measure of biological vulnerability, exacerbates these effects, with frail individuals showing heightened susceptibility to COVID-19 that scales non-linearly with their overall mortality risk (Hewitt et al., 2020; Shapiro et al., 2022).

Slurink et al. (2024), Xu et al. (2024), and Stivanello et al. (2022) found that mortality immediately after vaccination was significantly lower (Slurink et al. (2024) reported reductions of 70%, 50%, and 40% in weeks 1, 2, and 3, respectively), indicating self-selection of healthier individuals for vaccination. This missing mortality among the vaccinated leads to concentration effects in the unvaccinated, exacerbated by high vaccination rates. This could distort the relationship between COVID-19 mortality and non-COVID-19 mortality, as described in the Proportionality Hypothesis of Cairns et al. (2024), which posits that COVID- 19 mortality is approximately proportional to overall mortality within age groups and subgroups.

This study utilizes extensive data from the public database of the Office for National Statistics (ONS) in the United Kingdom, providing valuable insights into mortality patterns during the Delta and Omicron waves (Alessandria et al., 2025). The unique dataset includes five vaccination statuses (unvaccinated, one dose, two doses, three doses, four or more doses) and six age groups (18–39, 40–49, 50–59, 60–69, 70–79, 80–89, and 90+ years), based on the main dataset spanning 26 months (April 2021 to May 2023), supplemented with data from January 2021 to January 2022 to complete the picture for single-dose vaccinated and unvaccinated individuals. During this period, vaccination was administered four times with the same vaccine, including monthly data on COVID-19 mortality, all-cause mortality, and non-COVID-19 mortality, without artificial linking via matching. This offers a rare opportunity for detailed analysis of temporal mortality patterns within age groups, stratified by vaccination status, with a focus on the effects of repeated vaccinations. This study aims to investigate how selection bias and concentration effects influence the relationship between the relative risk of COVID-19 mortality ((RRcov)) and non-COVID-19 mortality ((RRnoncov)) within age groups. We hypothesize that this relationship is non-linear in the non-(further)- vaccinated subgroups, where a concentration of rapidly dying individuals and those recently infected has a disproportionate impact.

This relationship is modeled using a power function ((RRcov ∝ (RRnoncov)^a^)), as an extension of the linear proportionality from Cairns et al. (2024). Using the unique dataset, we can robustly test this non-linear relationship over a long follow-up period with repeated vaccinations, validating the optimal a-values through visual analysis of the vaccine effectiveness (VE) curves, which yield more stable and logical patterns at the correct a-values.

Furthermore, vaccination status misclassification can lead to an artificially high and prolonged post-vaccination effect in the data, as well as an apparently negative VE in the initial months, underscoring the need for careful correction in the analysis (Neil et al., 2022; Bakker et al., 2025; Fung et al., 2024; Agampodi et al., 2024).

## Methods

### Data Source

This study utilizes data from the public database of the Office for National Statistics (ONS) in the United Kingdom, which provides a unique dataset by supplying mortality rates for five vaccination statuses (unvaccinated, one dose, two doses, three doses, four or more doses) over a period of 29 months (January 2021 to May 2023), during which vaccination was administered four times with the same vaccine. This long follow-up period, including 26 months during the Delta and Omicron waves from April 2021 to May 2023 (Alessandria et al., 2025) with 3 additional months of annual data from January 2021 to March 2021 to complete the picture for single-dose vaccinated and unvaccinated individuals, enables an exceptional analysis of temporal mortality patterns and concentration effects in repeated vaccinations.

The dataset includes five vaccination statuses: unvaccinated (0 doses), one dose, two doses, three doses, and four or more doses. Mortality rates are stratified by six age groups: 18–39, 40–49, 50–59, 60–69, 70–79, 80–89, and 90+ years. Monthly data are available for COVID-19 mortality (Scov), non-COVID-19 mortality (Snoncov), and all-cause mortality (Sallcause). Age-standardized rates were used as reported in the ONS data; no exclusions were applied except for inherent dataset limitations (e.g., missing values). To make the trend clearly visible for each month, all age-standardized data with a number of deaths >3 were included. Data with <3 deaths were not included because no age-standardized data are provided by ONS for these. Data were downloaded from ONS (2023; 2022). The dataset names are: “All data relating to ‘Age-standardised mortality rates for deaths by vaccination status, England: deaths occurring between 1 April 2021 and 31 May 2023’” (Publication date: 25 August 2023) and “All data relating to ‘Age-standardised mortality rates for deaths by vaccination status, England: deaths occurring between 1 January 2021 and 31 May 2022’” (Publication date: 6 July 2022). Calculations were performed in Microsoft Excel. Raw data are available upon request. No individual patient data were used; the analysis is on an aggregated level, in accordance with UK data protection regulations – no IRB approval was required. This unique dataset enables a detailed and robust analysis of temporal mortality patterns within age groups, stratified by vaccination status, without artificial linking via matching.

### Hypothesis and Model

The hypothesis posits that the relative risk of COVID-19 mortality (RRcovi) is proportional to the relative risk of non-COVID-19 mortality (RRnoncov_i_) within a subgroup (i) (defined by vaccination status within an age group) according to a non-linear power function:

*RRvcov_i_ k*(RRnoncov_i_)^a^*
where (k) is a scaling constant and (a) is the exponent indicating the degree of non-linearity. A value of a > 1 points to a non-linear increase in (RRcovi), as expected due to selection bias in non-(further)-vaccinated groups, where concentration of rapidly dying individuals and those recently infected has a disproportionate impact. This non-linearity, potentially approaching quadratic dependence (a ≈ 2) in older age groups, likely arises from frailty among concentrated vulnerable individuals, as frailty disproportionately amplifies susceptibility to COVID-19 mortality compared to average non-COVID-19 causes of death (Shapiro et al., 2022; Hewitt et al., 2020). Relative risks are calculated relative to the unvaccinated group (0x) within the same age group, where

*RRcov_i_ = Scov_i_ / Scov_0x_ and RRnoncov_i_ = Snoncov_i_ / Snoncov_0x_*

By using relative risks, variations in infection rates are eliminated, as both vaccinated and unvaccinated groups are exposed to the same infection rates at the same time. This advantage makes the analysis more robust against external factors such as pandemic waves.

To quantify the impact of vaccination status, vaccine efficiency (VE) is calculated as:

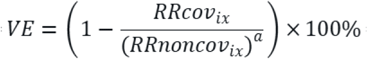

where RRcovix and RRnoncovix are the relative risks for the group with (i) doses, normalized relative to the unvaccinated (0x). This definition measures the relative reduction in COVID-19 mortality after correction for selection bias via (RRnoncovix)^a^. For the linear proportionality hypothesis (a = 1), VE is directly based on RRcov_ix_ / RRnoncov_ix_. For the non-linear hypothesis (a ≠ 1), (a) corrects for the non-linear impact of selection bias. VE is plotted against time for different a-values (e.g., a = 0, 1, 1.5, 2, 2.5, 3) to evaluate stability and logical patterns.

### Analysis Procedure

The optimal a-value yields stable VE curves with logical patterns, indicating robust correction for selection bias. This is determined through visual analysis of the VE curves based on the following criteria:

- Is there a logical pattern visible with increasing vaccine effectiveness over time for the single-dose vaccinated (1x)? For incorrect values, an odd illogical dip is visible in this group in the first months.
- Is the vaccine effectiveness always positive before February 2022? Additionally, the VE value at the start of the 1x vaccinated group is noted.
- Is the VE value always better than or equal to −50%?
- Are there indications of overcorrection (often clearly visible with the a=3 curve), for example, apparent extremely high VE values approaching 100%? Or indications of undercorrection where underlying patterns of mortality variation in the group are still visible.
- Do the regression lines approximately coincide or are they strongly different? What is the slope of the regression lines.

Additionally, the concept of the concentration factor (CF_i_) is introduced as a value indicating how strongly a non- or no-longer-vaccinated group is concentrated. Individuals who continue with vaccination (group nx (with n>i)) leave the “rapidly dying” or recently COVID-infected behind in the no-longer-to-be-vaccinated group (i), leading to mortality peaks as analyzed in the results. The concentration factor can be calculated as:

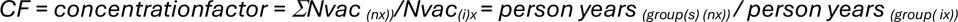

This demographic ratio (CF) quantifies the maximum potential for concentration by indicating how much the group is reduced in size, providing an upper limit for bias amplification; the observed RRnoncov elevation (e.g., up to 6 in 70-79-year-olds, Appendix II-C) is typically lower, depending on the proportion of individuals aware of their impending mortality, which may vary by age group. Nonetheless, CF indicates how strongly misclassification and frailty can influence outcomes.

This approach ensures a robust, bias-corrected estimation of VE. Due to data variability, the analysis relies on visual inspection rather than inferential statistics. Future analyses could incorporate statistical methods, such as non-linear regression or goodness-of-fit tests, to validate optimal a-values more objectively.

## Results and Discussion

### Vaccination Status and Concentration Factors

In Appendix I-A, person-years by vaccination status are presented as percentage values in bar charts for all age groups and vaccination statuses (0x, 1x, 2x, 3x, 4x). The oldest age groups (80–89 and 90+ years) were vaccinated first, followed by younger groups. The first vaccinations began in January 2021, with the second dose after three months, the third after six months, and the fourth dose after another six months for most groups. Vaccination rates were very high in the oldest age groups, with only 2–2.5% unvaccinated and 0.2– 0.3% single-dose vaccinated, leading to high concentration factors (see Appendix I-B). Appendix I-B shows the ratio of the i-times vaccinated group versus the (i-1)-times vaccinated group (i.e., the no-longer- vaccinated group) as a function of time. Numerical values for maximum ratios per age group and vaccination status are included in the table in Appendix II-B2.

### Non-COVID-19 Mortality and Relative Risks

In Appendix II-A, the age-standardized non-COVID-19 mortality rate (Snoncov) for the unvaccinated group is plotted against time for all age groups. For the unvaccinated (0x) and single-dose vaccinated (1x) groups in the three oldest age categories (70–79, 80–89, and 90+ years), non-COVID-19 mortality is shown in Appendix II-B, including three additional months before April 2021 to provide a complete overview. At the start of each vaccination round, a clear peak in non-COVID-19 mortality is visible in the group that is not or no longer vaccinated. The height of this peak correlates with the concentration factor (see table in Appendix II-B) but is also influenced by the vaccination rate in the group, as higher uptake amplifies concentration effects. A clear peak in non-COVID-19 mortality is visible at each vaccination, particularly in the single-dose vaccinated, who experience the highest concentration factor (see table in Appendix I-B) because at least two doses were the norm.

In Appendix II-C, the relative non-COVID-19 mortality (RRnoncov) is plotted against time, showing that mortality in the single-dose vaccinated group of 70–79 years is increased by a factor of 6 due to extreme concentration; in the 80-89 year olds, the relative risk value (RRnoncov1x) is 5, and in the 90+ group, 3.5. This effect is most extreme in the single-dose vaccinated (see Appendix II-B and II-C), who form a transition group between unvaccinated and multiple-dose vaccinated. Furthermore, it is clear that non-COVID-19 mortality in the group that is revaccinated starts significantly lower than 1.0 due to shedding of the weakest individuals. Only when this group itself is concentrated—i.e., when a new group is vaccinated—does mortality in this group increase due to concentration occurring here. This pattern repeats in the 2x, 3x, and 4x vaccinated, with the effects becoming progressively smaller.

In Appendix II-B2, the concentration factor decreases for the 3rd and 4th vaccinations, especially in younger age groups, due to lower vaccination uptake. Additionally, the proportion of predictable mortality is lower in younger individuals, so concentration effects are expected to have a smaller impact in those groups anyway.

In the younger age groups (18-39 and 40-49 years), the 3x vaccinated are hardly concentrated because few people in these groups get a 4th vaccination. Furthermore, the relative risk of non-COVID mortality (RRnoncov) for the 4x vaccinated shows an anomalous pattern in the first months. The RRnoncov values often start high because during that period, a smaller part of the population was vaccinated, often only the weaker part that just qualified for vaccination. Only after a larger part of the population is vaccinated does the concentration effect become visible in an increase in RRnoncov among the no-longer-vaccinated and a significant decrease in RRnoncov for the four-times vaccinated, who then leave the rapidly dying behind and concentrated in the three-times vaccinated group.

### COVID-19 Mortality

In Appendix III-A, COVID-19 mortality (Scov) for all groups is plotted on the same scale, showing that total COVID-19 mortality is mainly determined by the age groups 70–79, 80–89, and 90+ years, with a small contribution from the 60–69 year olds. The contribution of younger groups (<60 years) is virtually nil. In Appendix III-B, COVID-19 mortality per age group is plotted on a custom scale, making differences clearer. In January 2022, a peak in COVID-19 mortality is visible for the unvaccinated (0x), single-dose (1x), and double-dose (2x) vaccinated, likely due to the Omicron wave, followed by a strong decline in COVID mortality. Data after February 2022 and before July 2021 therefore show larger measurement errors, limiting the reliability of these periods.

At first glance, it seems strange that 0x, 1x, and even 2x have virtually the same COVID mortality peak around December 2021 - January 2022. The vaccine seems not to work, but this is only apparent because the underlying groups differ greatly in mortality. This becomes clear further on when we look at vaccine effectiveness.

### Vaccine Effectiveness

In Appendix IV-I, the VE curves with a=1, 1.5 and 2 are plotted for age groups, providing the core visualization around optimal a-values to highlight stability and logical patterns. These form the primary analysis, as they focus on the values that best correct for non-linearity. Appendix IV-I1 shows this for age groups ≥70–79 years, and Appendix IV-I2 for ≤60–69 years.

In Appendix V-A to Appendix V-C, the VE curves for the three oldest age groups (70+) are plotted in a way that highlights key patterns, as these groups contribute most to overall COVID-19 mortality and exhibit the strongest non-linear effects. These visualizations form the core of the analysis, showing VE for a=0, 1, 1.5, 2, 2.5, and 3 to demonstrate the impact of the a-value. For completeness, Appendix VI-A to Appendix VI-G provides the VE curves for a = 0, 1, 1.5, 2, 2.5, and 3 for all age groups, allowing a broader comparison of the total pattern of VE as a function of time across all groups.

In Appendix VI-H1-4, the visual analysis of the different graphs is presented in an overview. From the visual analysis of the VE graphs (according to criteria in Methods), values of a = 1.5 to 2 emerge for the 80–89-year group and 90+ groups, and 1.5 for the 70–79-year-olds. If negative VE is excluded, a lies at 1.5. If negative VE is allowed (because vaccination may initially lead to reduced resistance), a value of 2 for the oldest two age groups is more likely. However, the results contain too large an error to designate either value as the correct one. Thus, a = 1.5-2 is optimal for 80+ and for 70-79 years, a-value of 1.5 suffices, while for the younger age groups, an a-value of 1 to 1.5 makes little difference. The a-value for age groups <70 years probably matters much less in these younger age groups because the concentration factors were lower. At a = 1, all curves in the younger groups therefore show approximate overlap. For the 18–39-year-olds, the conclusion is limited because there were very few data points, but they seemed to show the same VE trends.

The level of VE shows a logical pattern at optimal a-values, with levels stabilizing around 60-90% in a broad band of noise, caused by the relatively large error in COVID-19 mortality in periods with little COVID. There is no large negative VE visible, and the curves for the single-dose vaccinated group show no strange dip in the first months, indicating consistent protection after the initial period. Also, overcorrection (visible by a reversed time trend) is avoided. But the most striking is that the time trend at optimal a-values seems to have completely disappeared: more than 1 vaccination does not seem to further improve VE.

## Discussion

### Interpretation of Results

The results show that the unvaccinated groups, especially in the oldest age groups (70-79, 80–89 and 90+ years), exhibit a high concentration of rapidly dying individuals and those recently infected, as evidenced by the increased non-COVID-19 mortality and concentration factors (Appendix I-B). These peaks in non-COVID-19 mortality suggest that self-selection and the subsequent concentration effects strongly influence mortality in non-(further)-vaccinated groups, particularly in the single-dose vaccinated who experience extremely high concentration factors.

COVID-19 mortality is largely determined by the oldest age groups, with a peak in January 2022 due to the Omicron wave. The repeatedly low mortality in newly vaccinated groups corresponds to self-selection of healthier individuals, as found by Slurink et al. (2024). Vaccine efficiency is high immediately after vaccination but declines rapidly at a=1, while the non-linear model (RRcov ∝ (RRnoncov)^a^) yields more stable VE values, indicating disruption of the linear proportionality hypothesis of Cairns et al. (2024) by selection bias and underlying frailty.

The convergence of VE curves across vaccination statuses at optimal a-values (1.5–2), despite measurement errors in low-COVID periods (Appendices V-A–C and VI-H1-4), provides compelling evidence that underlying frailty disproportionately influences COVID-19 mortality more than previously recognized, or indicates systematic misclassification bias. Frailty amplifies COVID risks non-linearly in concentrated non-vaccinated groups, requiring higher a-value for correction (Shapiro et al., 2022; Hewitt et al., 2020). Alternatively, misclassification artificially elevates mortality in lower-dose groups, skewing curves until non-linear adjustment stabilizes VE (Neil et al., 2022; Bakker et al., 2025). This overlap suggests that observed waning VE in prior studies may stem from uncorrected biases rather than true decline.

### Comparison with Literature

The dominance of older groups in COVID-19 mortality supports the proportionality hypothesis, but the observed peaks in non-COVID-19 mortality in unvaccinated individuals suggest that selection bias disrupts this, with lower effects in younger groups due to lower concentration factors. This aligns with Alessandria et al. (2025), who found rising SMRs and RRs in vaccinated groups due to waning effectiveness and bias. The power dependence (a ≈ 1.5 to 2) explains the decline in survival among unvaccinated, as in Kaplan-Meier curves from Bakker et al. (2025). The extreme concentration in older groups emphasizes correction for bias, as suggested by Alessandria et al. (2025).

The disappearance of the strong time dependence of Vaccine Effectiveness after correction for power dependence seems to indicate that earlier observations of a strongly waning Vaccine Effectiveness, as observed by Tartof et al. (2021) and Goldberg et al. (2021), may be based on insufficient correction for the Healthy Vaccinee effect.

While Cairns et al. (2024) find linearity in aggregate data over all ages, our within-age-group analysis reveals non-linearity driven by frailty and vaccination biases, extending their hypothesis to finer sub-groups where proportional assumptions break down.

Moreover, misclassification is suspected in individuals who die shortly after vaccination, where death registration may be entered into the registry before the vaccination itself (Neil et al., 2022). Even if this occurs in only 1% of cases among vaccinated individuals, it can – due to the enormous concentration effects where the vast majority of the population is further vaccinated and only “rapidly dying” and recently COVID-infected remain – significantly increase mortality in the unvaccinated group. This affects both COVID-19 and non-COVID-19 mortality proportionally. By modeling the relationship with a power function (RRcov ∝ (RRnoncov)^a), this effect is incorporated into the relationship, independent of the presence of misclassification.

### Implications of Selection Bias

The analysis suggests that the high vaccination rate in the UK (>95% for ≥70-year-olds) led to unnecessary vaccinations due to inadequate correction for the healthy vaccinee effect (HVE). Matching was insufficient, as demonstrated by Bakker et al. (2025). Using ONS data over 29 months, the relationship proved non-linear in older individuals (a ≈1.5-2), explaining the decline in survival among unvaccinated. At a=1.5, one vaccination appears sufficient for optimal protection; multiple doses provide no improvement. This has implications for strategies: incorporate better HVE corrections to avoid overvaccination, as emphasized by Alessandria et al. (2025). Targeting frail populations could optimize future policies. These findings align with studies highlighting the need for robust bias correction in observational data to inform vaccination policies (Fung et al., 2024; Obel et al., 2024).

### Limitations

The analysis is limited by measurement errors in the ONS data, especially due to low numbers of COVID-19 deaths in subgroups (particularly after February 2022 and before July 2021) – which is clearly visible in the noise in the data during this period. For missing infection rates, correction was made by looking at relative risks over time. Misclassification can distort results (Bakker et al., 2025; Neil et al., 2022, Fung et al., 2024) The visual inspection of VE curves in Appendices IV-A to IV-G determines the optimal a-values, which may limit robustness without additional statistical validation. Future research should incorporate quantitative model fits (e.g., goodness-of-fit tests) and individual-level data to reduce subjectivity and address potential confounding, such as health-seeking behavior or prior infection history, as highlighted in observational studies (Obel et al., 2024; Agampodi et al., 2024).

### Conclusions

For older age groups (70-79 years, 80–89 years, and the 90+ age group), the relationship between the relative risk of COVID-19 mortality ((RRcov)) and non-COVID-19 mortality ((RRnoncov))is non-linear, with optimal exponents of a-values around 1.5 to 2, based on visual analysis of VE curves (Appendices IV-A to IV-G). These findings, enabled by the unique ONS dataset tracking five vaccination statuses over 29 months with four vaccinations, confirm the robustness of the non-linear model and explain the enormous decline in survival among unvaccinated during the first vaccinations, as observed in Kaplan-Meier curves (Bakker et al., 2025).

For younger age groups (<70 years), a = 1 or 1.5 suffices for the relationship RRcov ∝ (RRnoncov)^a; a value between 1 or 1.5 has little impact on the overall outcomes of Vaccine Effectiveness.

With the obtained results according to the model ((RRcov∝(RRnoncov)^a^)), the second, third, and fourth vaccinations appear to provide no significant additional benefit, though further validation with statistical testing is needed.

The vaccine efficiency, defined as VE = (1 - RRcov (RRnoncov)^a^) × 100%, lies in the period until the end of 2021 between 60-90% for the oldest age groups (with a broad band due to the error in small numbers of COVID deaths) at optimal a-values, but declines after February 2022, possibly due to waning immunity or pandemic waves or because the virus has circulated in the unvaccinated groups with which it is compared.

The high mortality among non-further-vaccinated groups after vaccination moments is explained by self-selection and high concentration factors.

The non-linear model RRcov ∝ (RRnoncov)^a^ with relative risks normalized relative to unvaccinated, offers a more robust approach for mortality analysis by reducing the impact of vaccination status misclassification, as supported by the unique dataset and explains the extreme decline in survival of unvaccinated in the Kaplan-Meier curves of Bakker et al. (2025) and corrects for possible misclassification (Alessandria et al., 2025; Neil et al., 2022) by incorporating this effect into the non-linear relationship. Frailty provides a mechanistic explanation for this non-linearity, particularly in older cohorts, warranting targeted interventions.

## Data Availability

All data produced are available online at https://doi.org/10.6084/m9.figshare.29926580.v1

## Appendices

**Appendix I: Concentration ratio, person-years by vaccination status**

Appendix I-A: Person-years by vaccination status (Appendix A1: 3D Appendix A2: 2D)

Appendix I-B: Concentration ratio of i-times vaccinated group versus (i-1)-times vaccinated group (person-years/person-years)

Appendix II-B2: Maximum Concentrationfactor (CF) for different age groups and i-times vaccinated groups during vaccination

**Appendix II: Non-COVID-19 mortality (Snoncov, RRnoncov)**

Appendix II-A: Age-standardised non-COVID-19 mortality rate per 100.000 person-years (Snoncovi) for all age groups and vaccination statuses

Appendix II-B: non-COVID mortality rate (Snoncovi) for 70-79 years, 80-89 years and 90+ years age groups unvaccinated (0x) and once (1x) vaccinated-groups

Appendix II-C Relative Risk of Non-COVID-19 mortality (RRnoncovi) versus time

**Appendix III: Covid mortality (Scov)**

Appendix III-A: COVID-19 mortality (Scovi) for all age groups and vaccination statuses (uniform scale)

Appendix III-B: COVID-19 mortality (Scovi) for all age groups and vaccination statuses (custom scale)

**Appendix IVI-I: Vaccine Effectiveness with a=1, 1.5 and 2**

Appendix IVI-I1: Vaccine Effectiveness with a=1, 1.5 and 2 for Age groups ≥70–79 Years

Appendix IVI-I2: Vaccine Effectiveness with a=1, 1.5, and 2 for Age groups ≤60–69 Years

**Appendix V: Vaccine Effectiveness versus time for 70+ years groups**

Appendix V-A: Vaccine Effectiveness for the 90+ age group

Appendix V-B: Vaccine Effectiveness for the 80-89 age group

Appendix V-C: Vaccine Effectiveness for the 70-79 age group

**Appendix VI: Vaccine Effectiveness versus time**

Appendix VI-A: Vaccine Effectiveness for a=0, 1, 1.5, 2, 2.5 and 3 in the 90+ Age Group

Appendix VI-B: Vaccine Effectiveness for a=0, 1, 1.5, 2, 2.5 and 3 in the 80–89 Age Group

Appendix VI-C: Vaccine Effectiveness for a=0, 1, 1.5, 2, 2.5 and 3 in the 70–79 Age Group

Appendix VI-D: Vaccine Effectiveness for a=0, 1, 1.5, 2, 2.5 and 3 in the 60–69 Age Group

Appendix VI-E: Vaccine Effectiveness for a=0, 1, 1.5, 2, 2.5 and 3 in the 50–59 Age Group

Appendix VI-F: Vaccine Effectiveness for a=0, 1, 1.5, 2, 2.5 and 3 in the 40–49 Age Group

Appendix VI-G: Vaccine Effectiveness for a=0, 1, 1.5, 2, 2.5 and 3 in the 18–39 Age Group

**Appendix VI-H1-4: Analyses of Vaccine Effectiveness: Graphs for different a-values**

---

*Appendix I-A: Person years by vaccination status)*

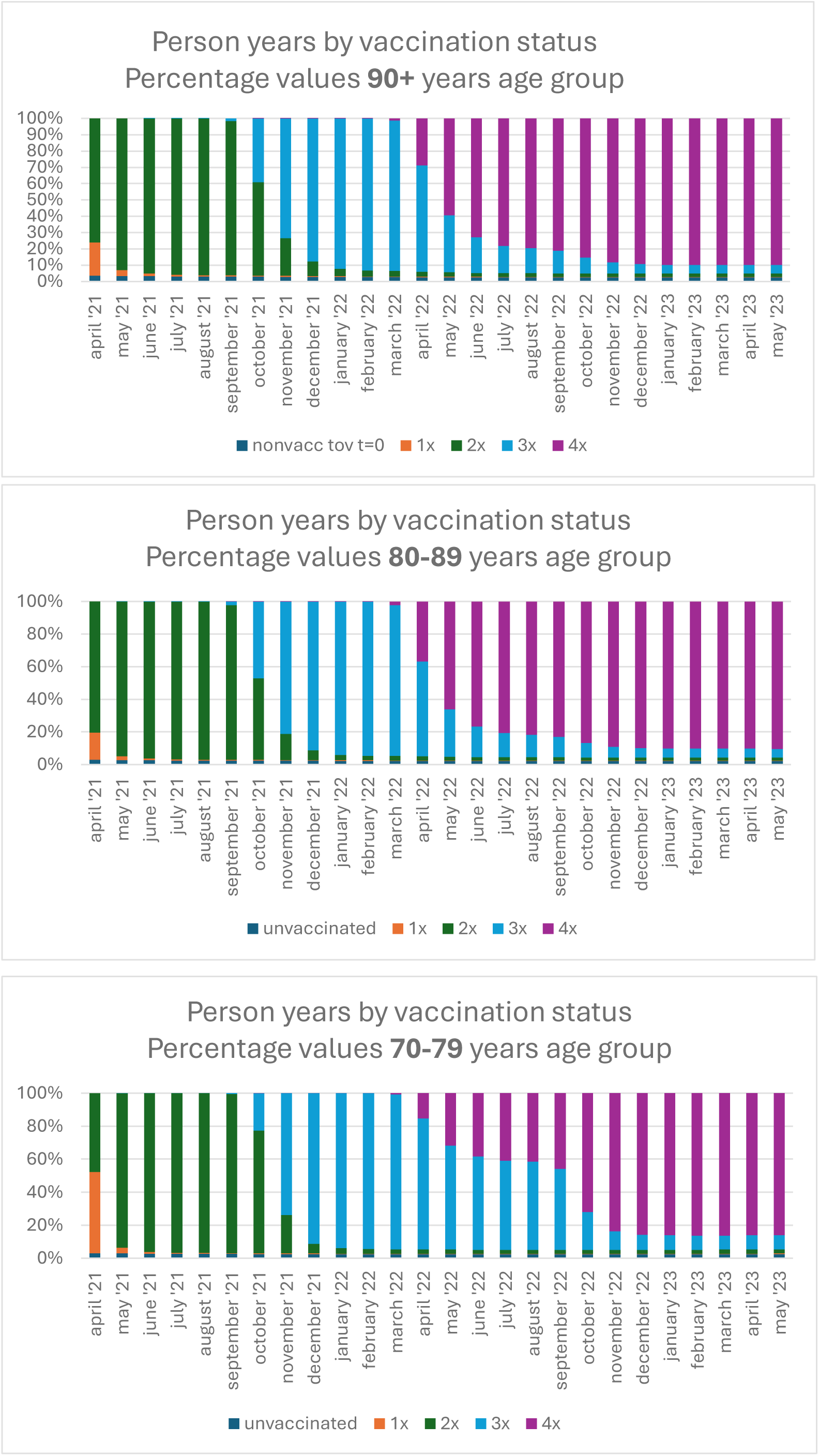

*Appendix IA: Person years by vaccination status (part 2)*

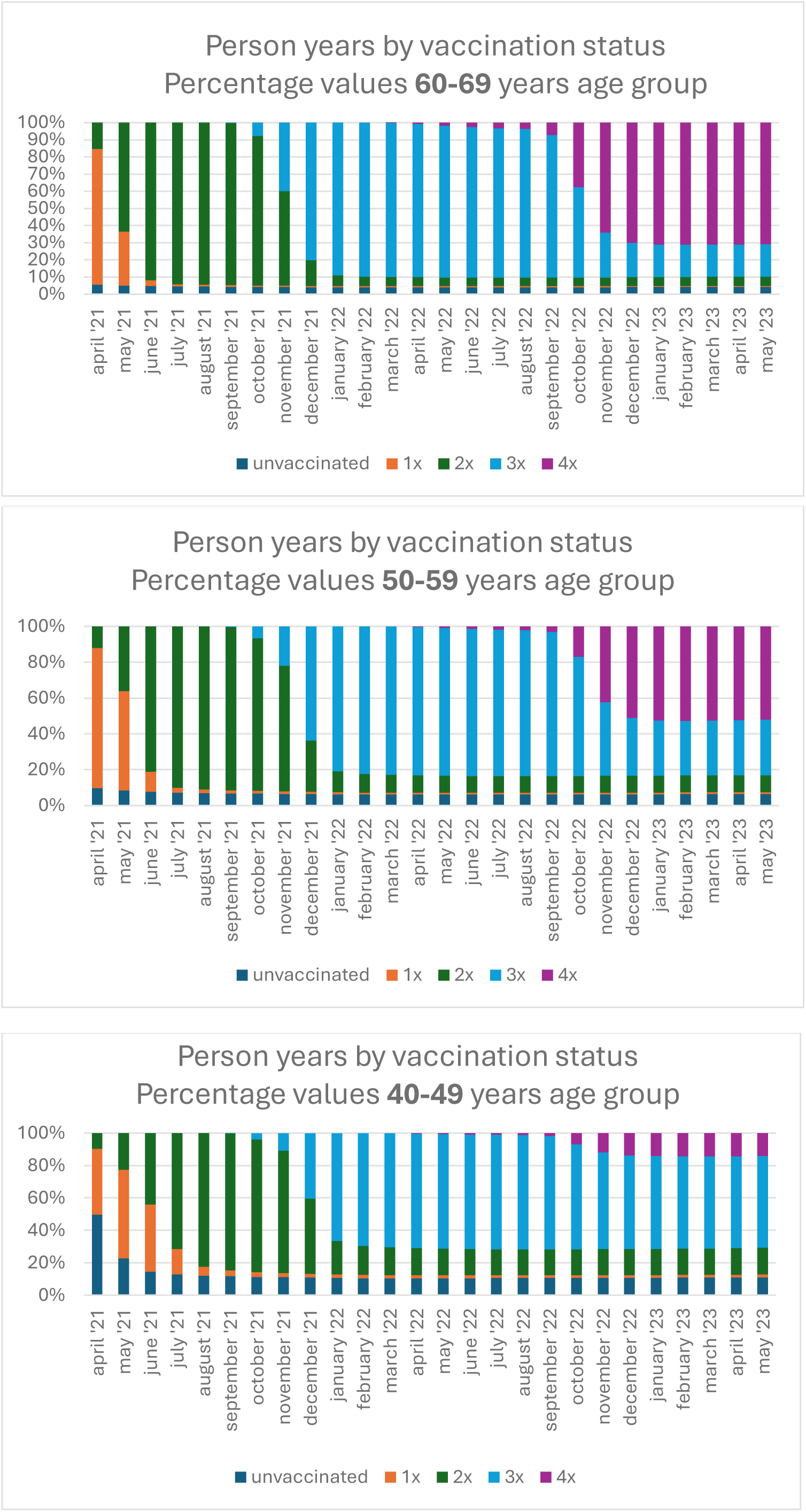

*Appendix IA: Person years by vaccination status (part 3)*

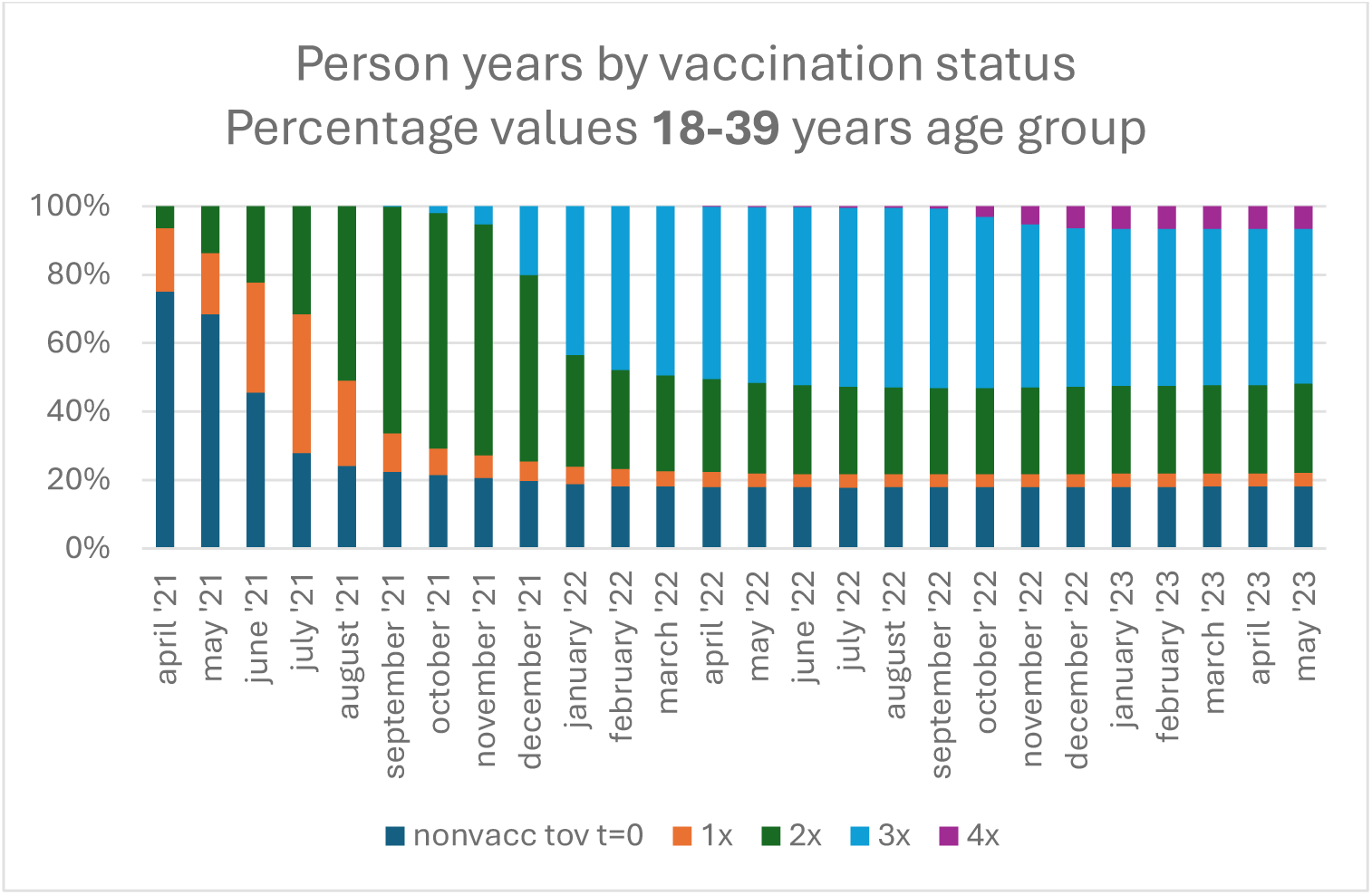

*Appendix I-B: Concentration ratio of i-times vaccinated group versus (i-1)-times vaccinated group (person-years/person-years)*

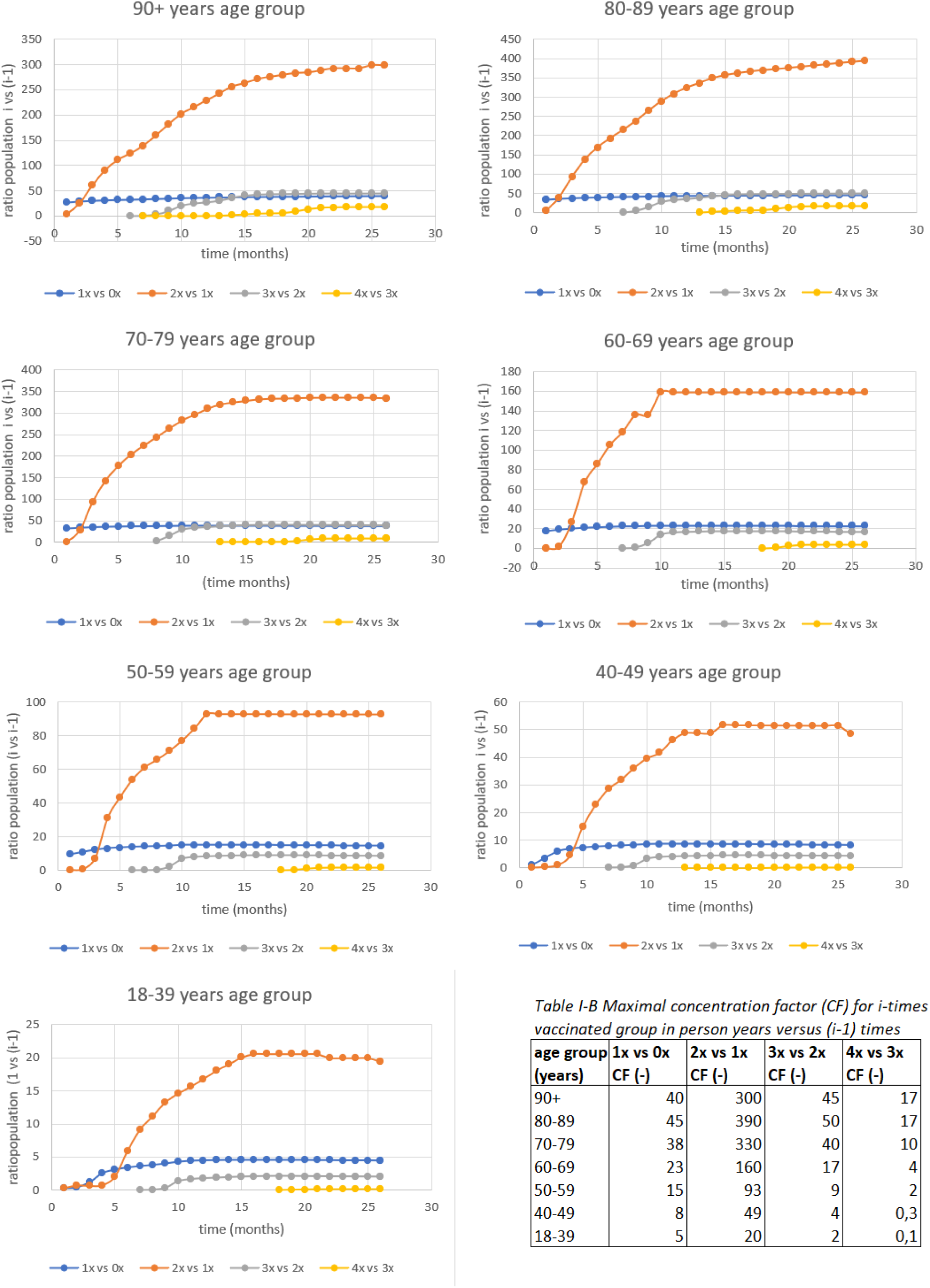

*Appendix II-A: Age-standardised Non-COVID-19 mortality rate per 100.000 person-years (Snoncovi) for all age groups and vaccination statuses*

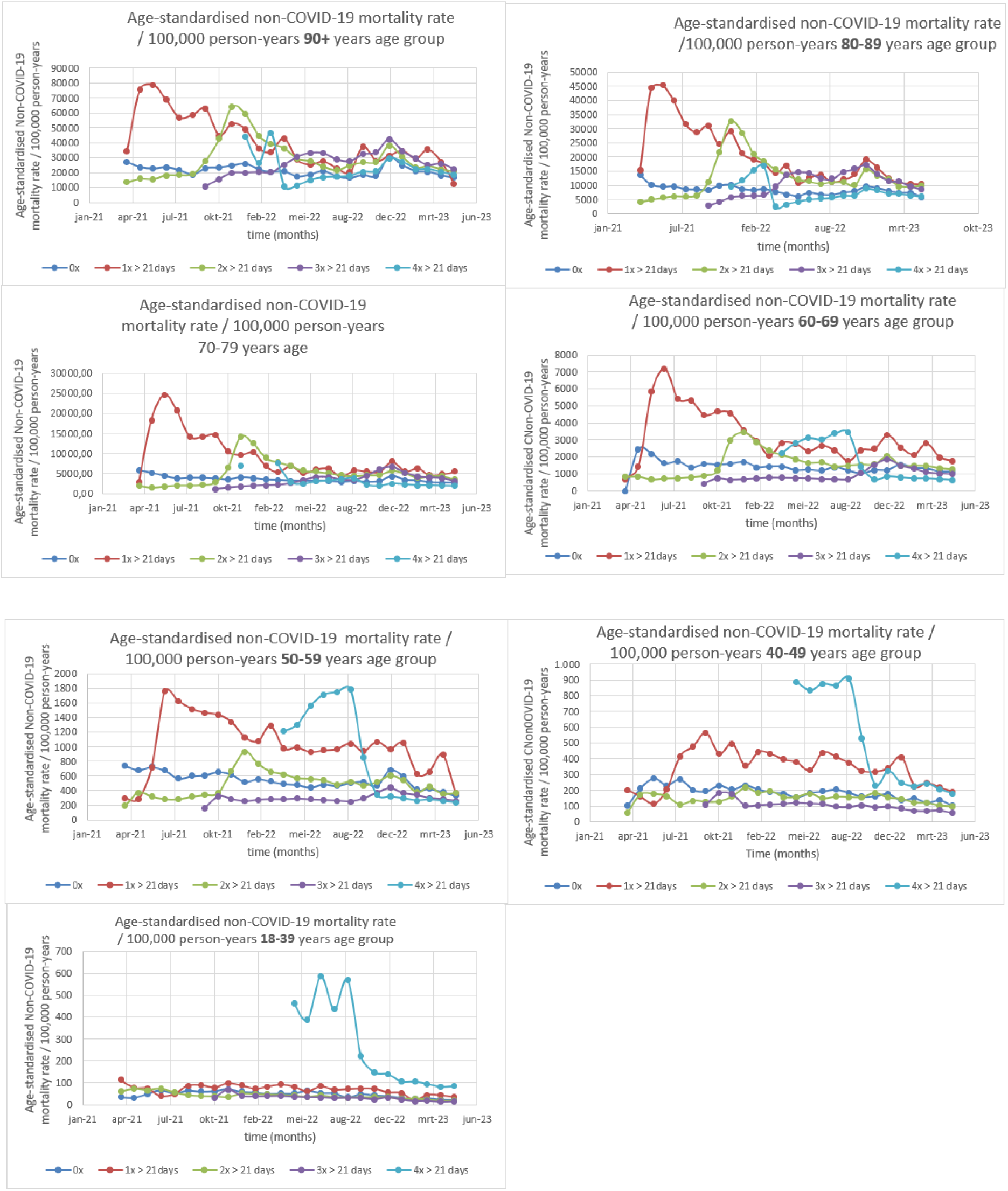

*Appendix II-B: non-COVID mortality rate (Snoncovi) for 70-79 years, 80-89 years and 90+ years age groups unvaccinated (0x) and once (1x) vaccinated-groups*

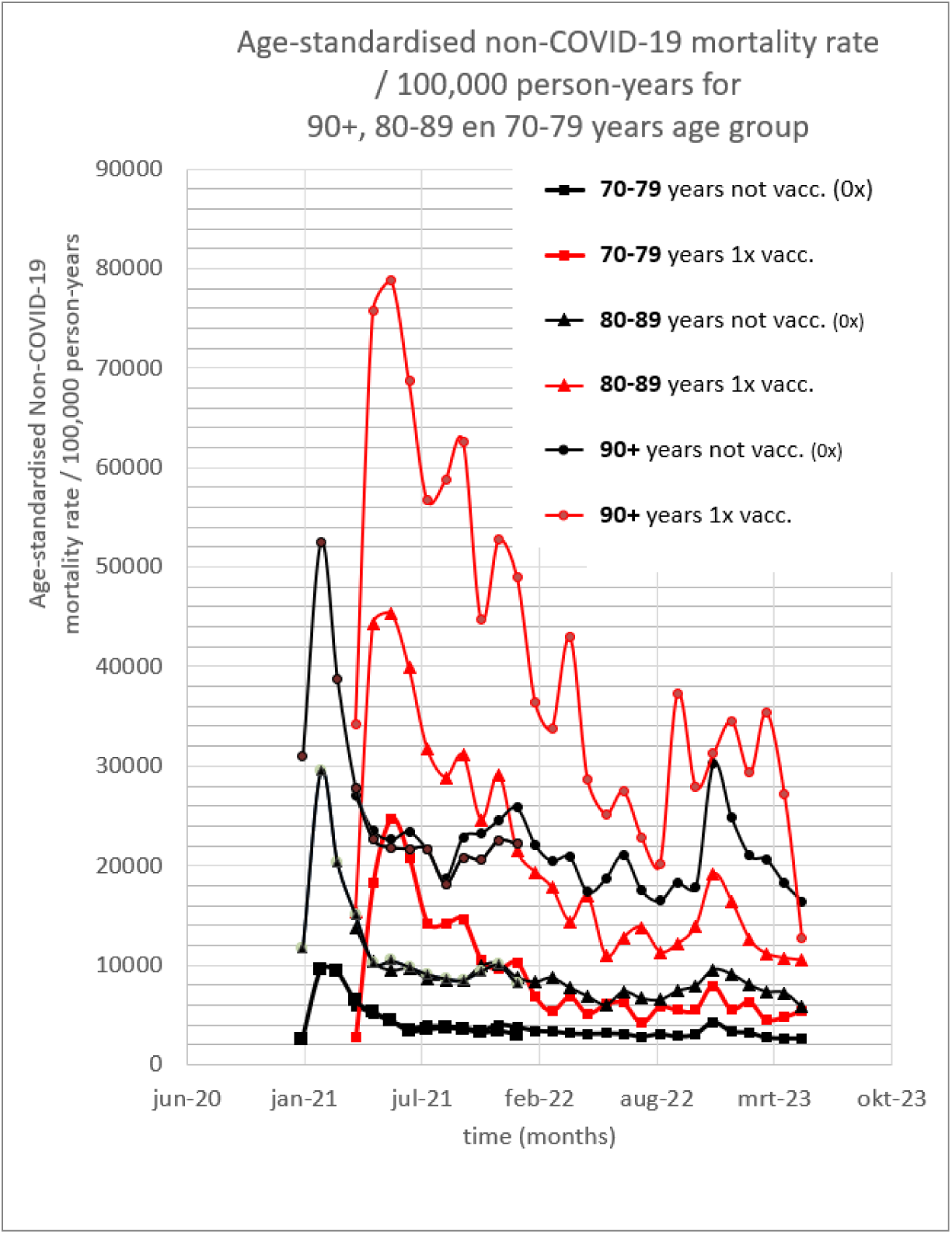

*Appendix II-C Relative Risk of non-COVID-19 mortality (RRnoncovi) versus time*

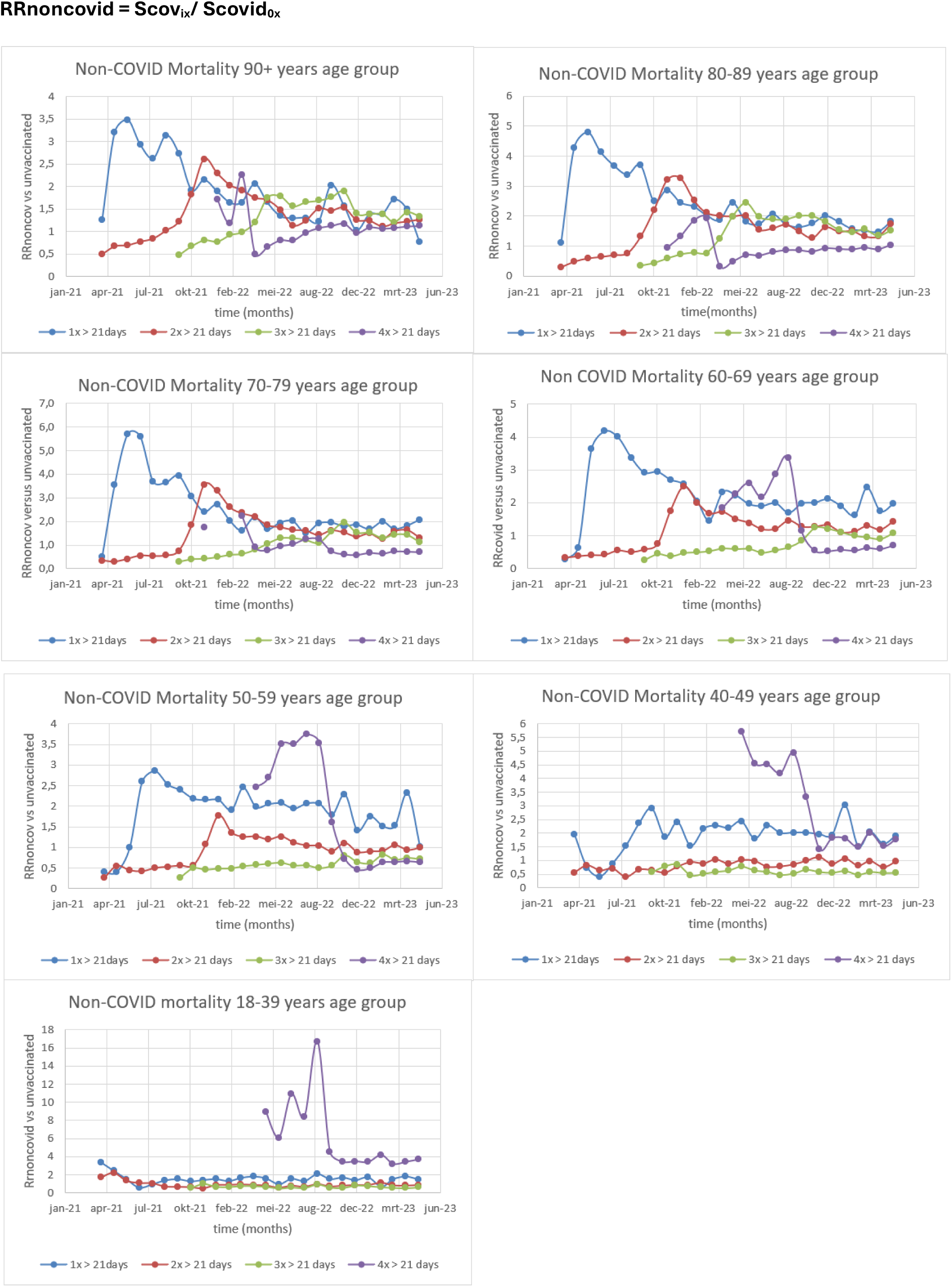

*Appendix III-A: COVID mortality (Scovi) for all age groups and vaccination statuses (uniform scale)*

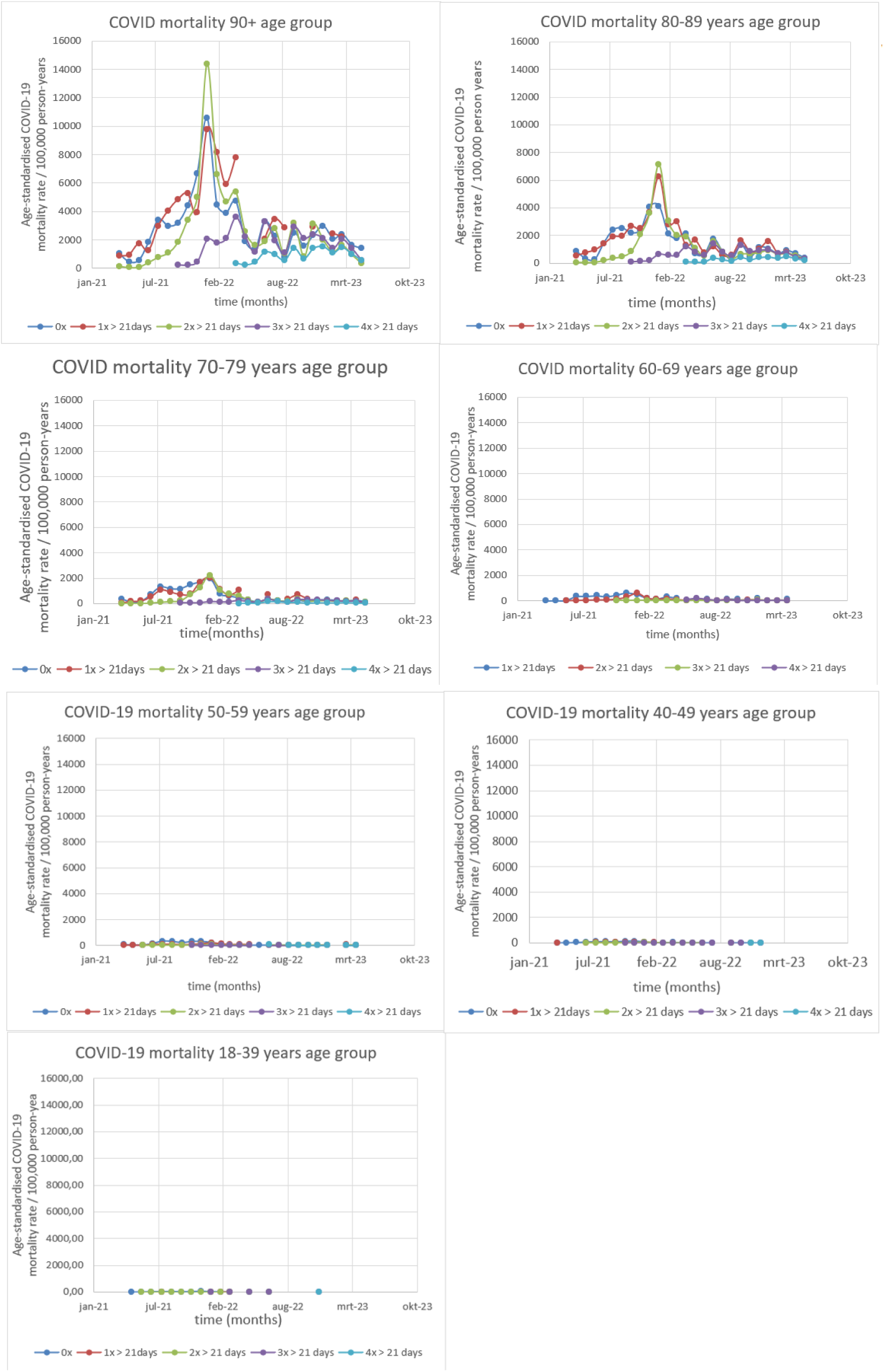

*Appendix III-B: COVID mortality (Scovi) for all age groups and vaccination statuses (custom scale)*

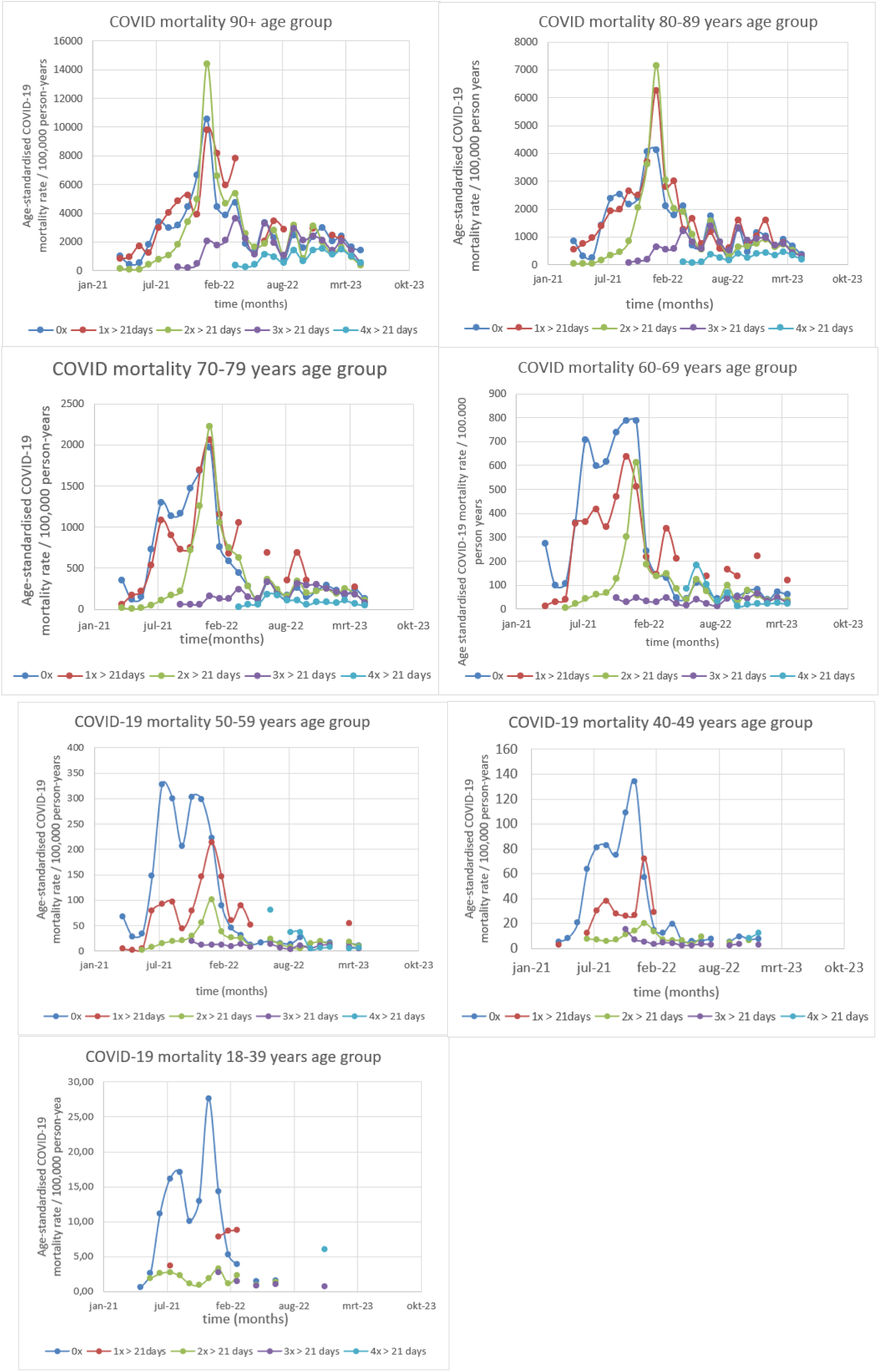

*Appendix IV-I1: Vaccine Efficiency with a =1*, *1,5 and 2 for all age groups*

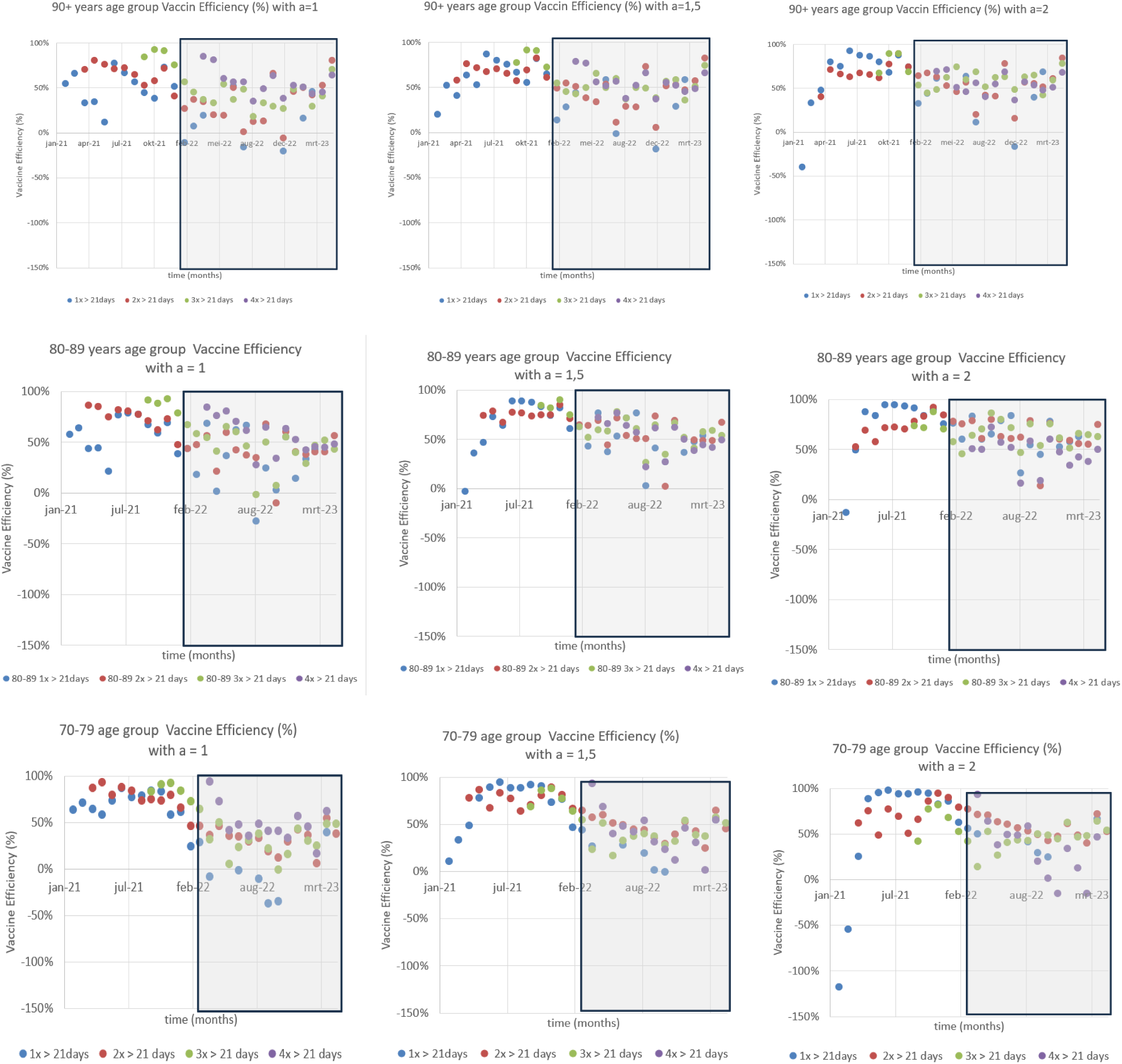

*Appendix IV-I2: Vaccine Efficiency with a =1*, *1,5 and 2 for all age groups (part 2)*

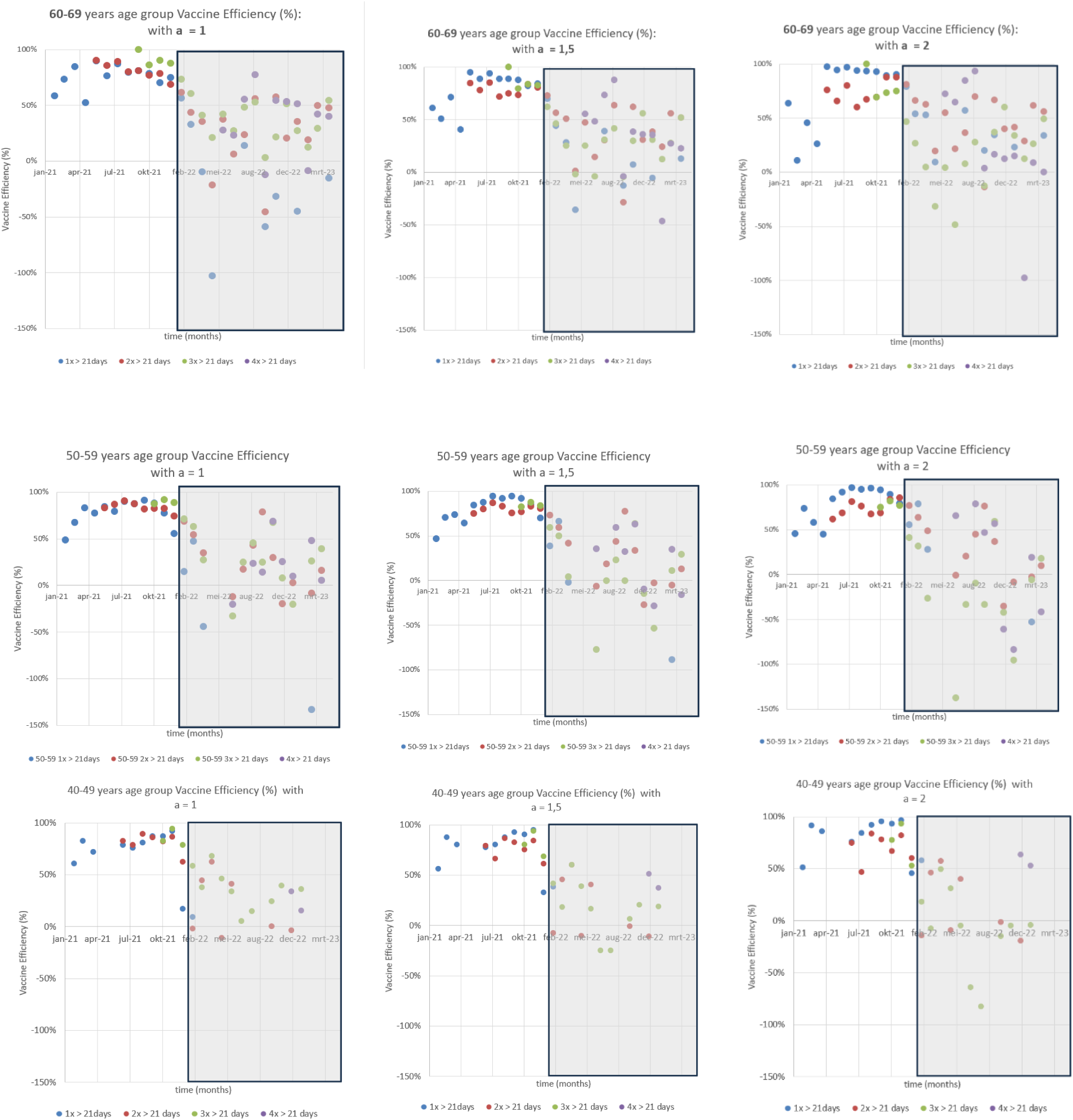

*Appendix IV-I3: Vaccine Efficiency with a =1*, *1,5 and 2 for all age groups (part 3)*

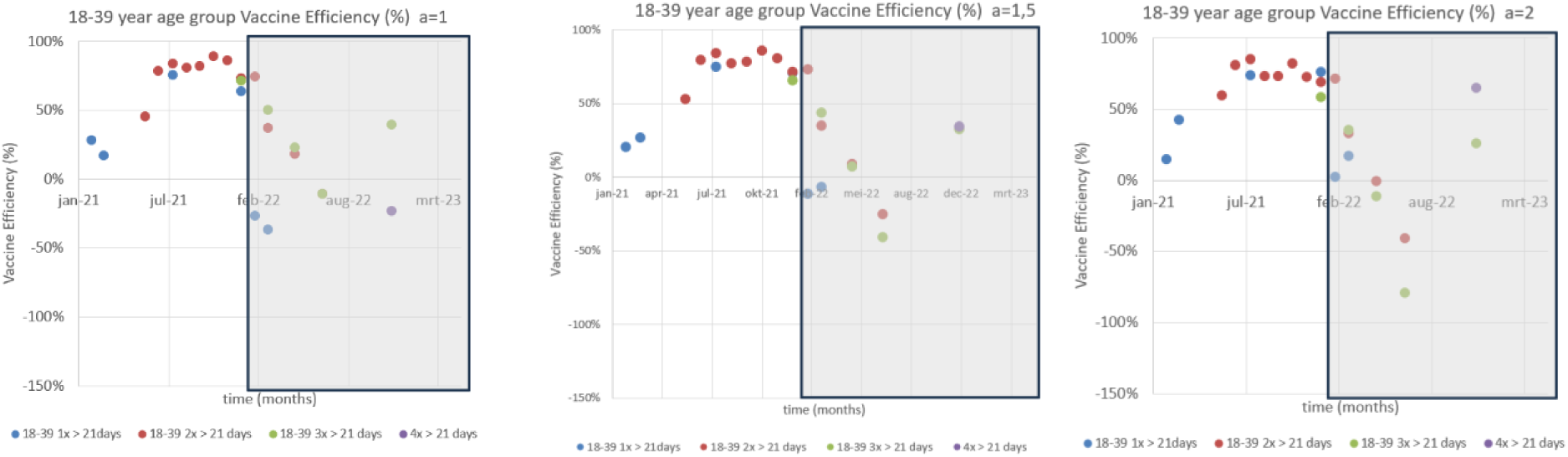

*Appendix V-A: Vaccine Effectiveness versus time: 90+ years age group for different a-values*

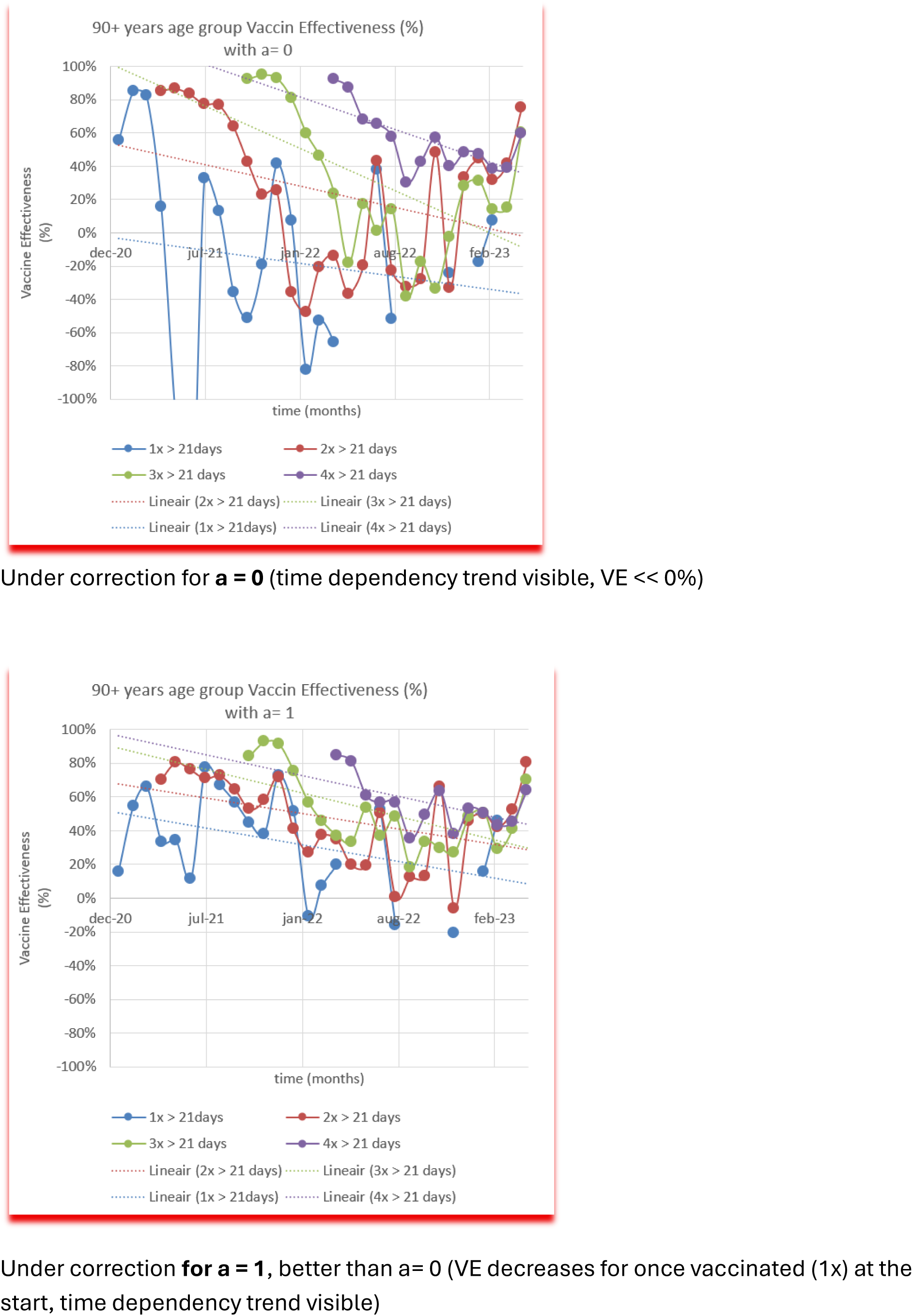

*Appendix V-A: Vaccine Effectiveness versus time: 90+ years age group for different a-values (part 2)*

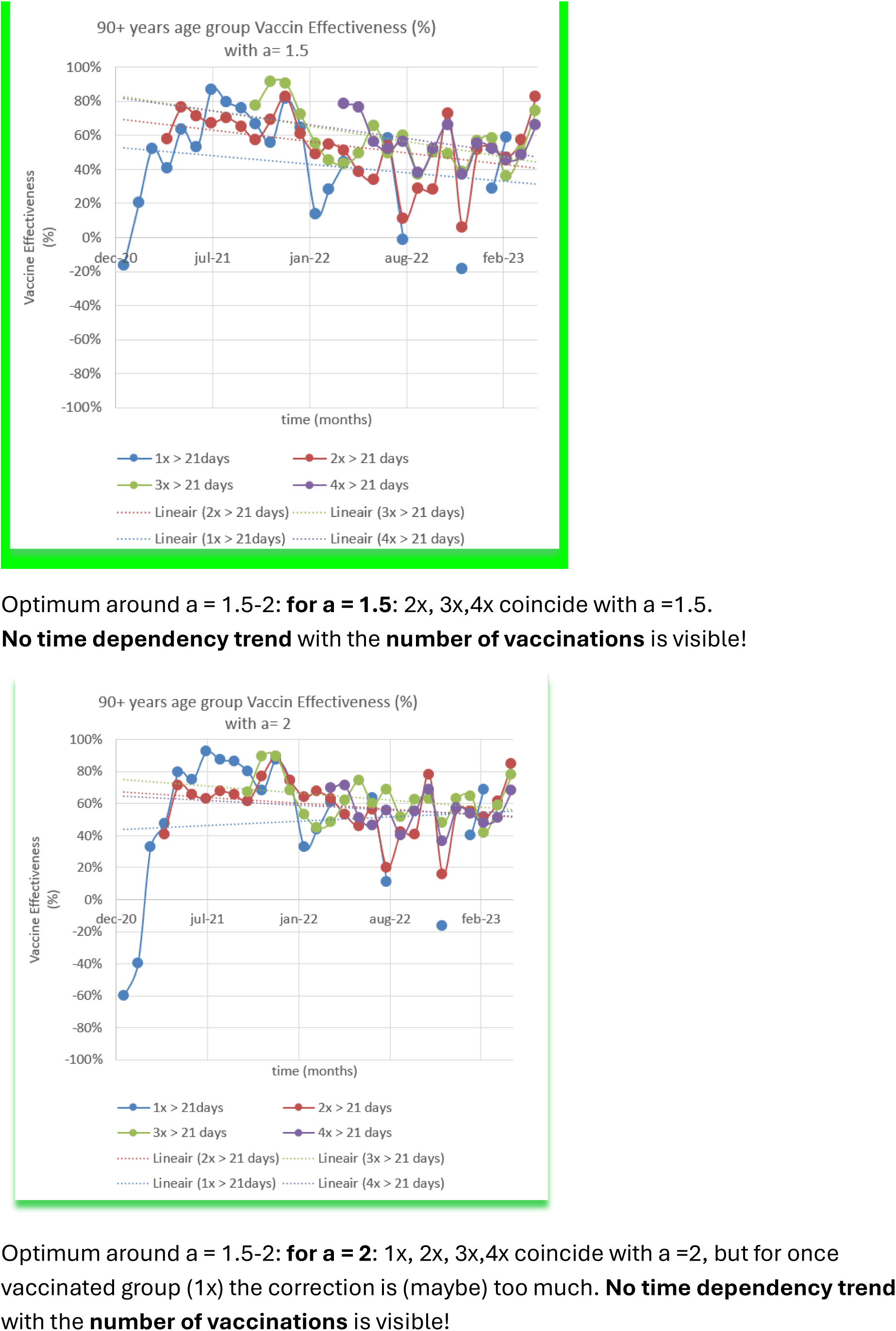

*Appendix V-A: Vaccine Effectiveness versus time: 90+ years age group for different a-values (part 3)*

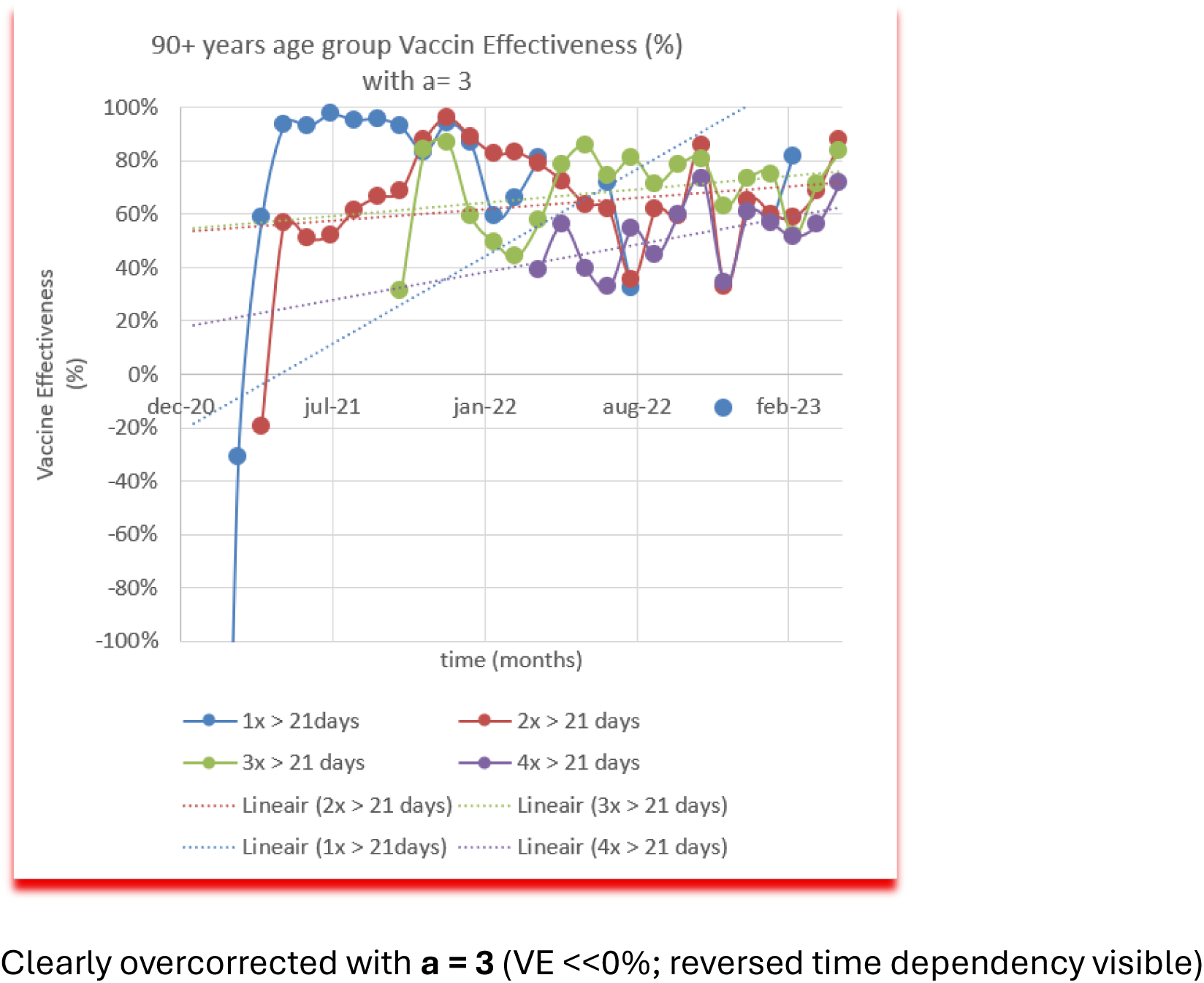

*Appendix V-B: Vaccine Effectiveness versus time: 80-89 years age group for different a-values (part 1)*

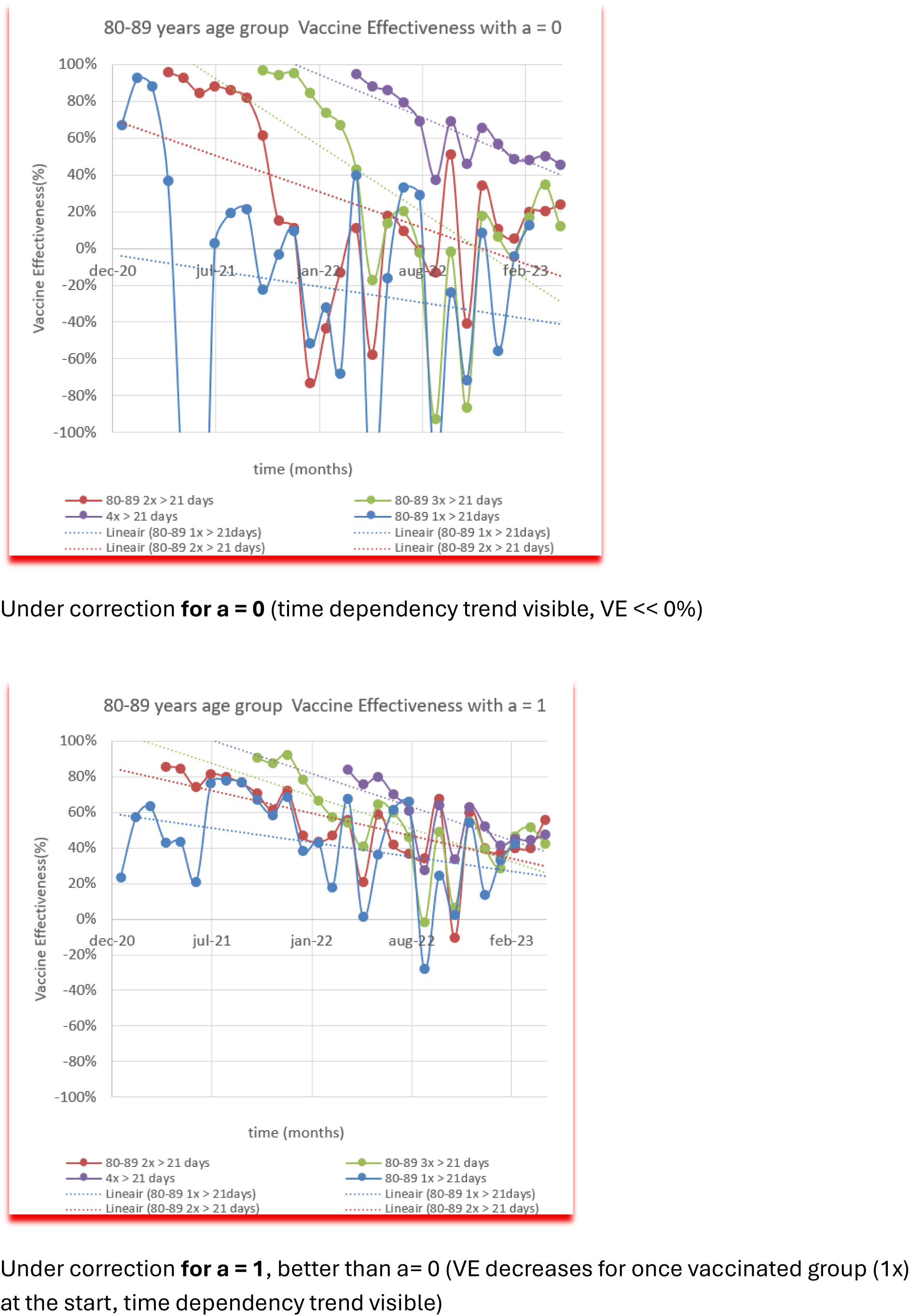

*Appendix V-B: Vaccine Effectiveness versus time: 80-89 years age group for different a-values (part 2)*

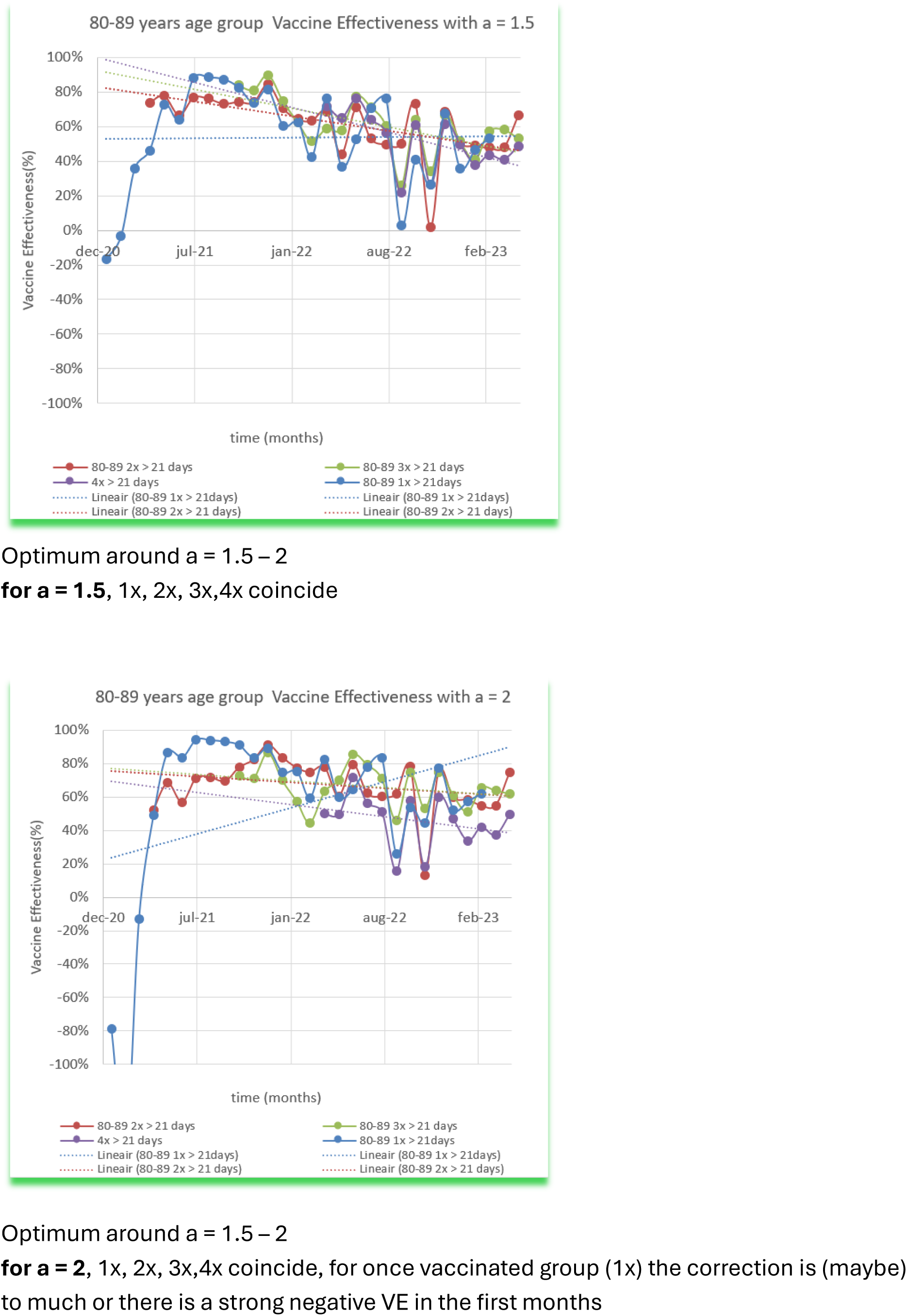

*Appendix V-B: Vaccine Effectiveness versus time: 80-89 years age group for different a-values (part 3)*

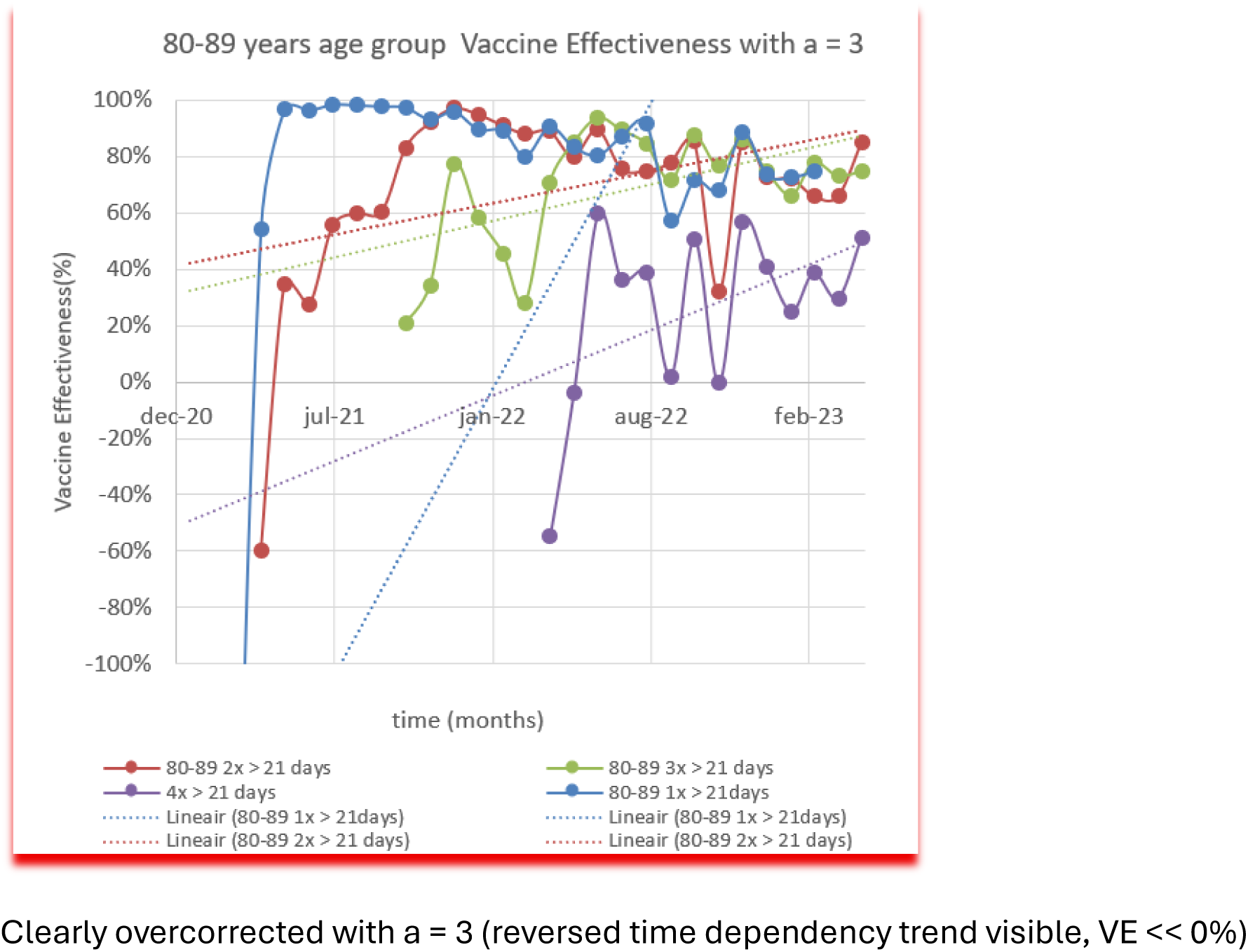

*Appendix V-C: Vaccine Effectiveness versus time: 70-79 years age group for different a-values (part 1)*

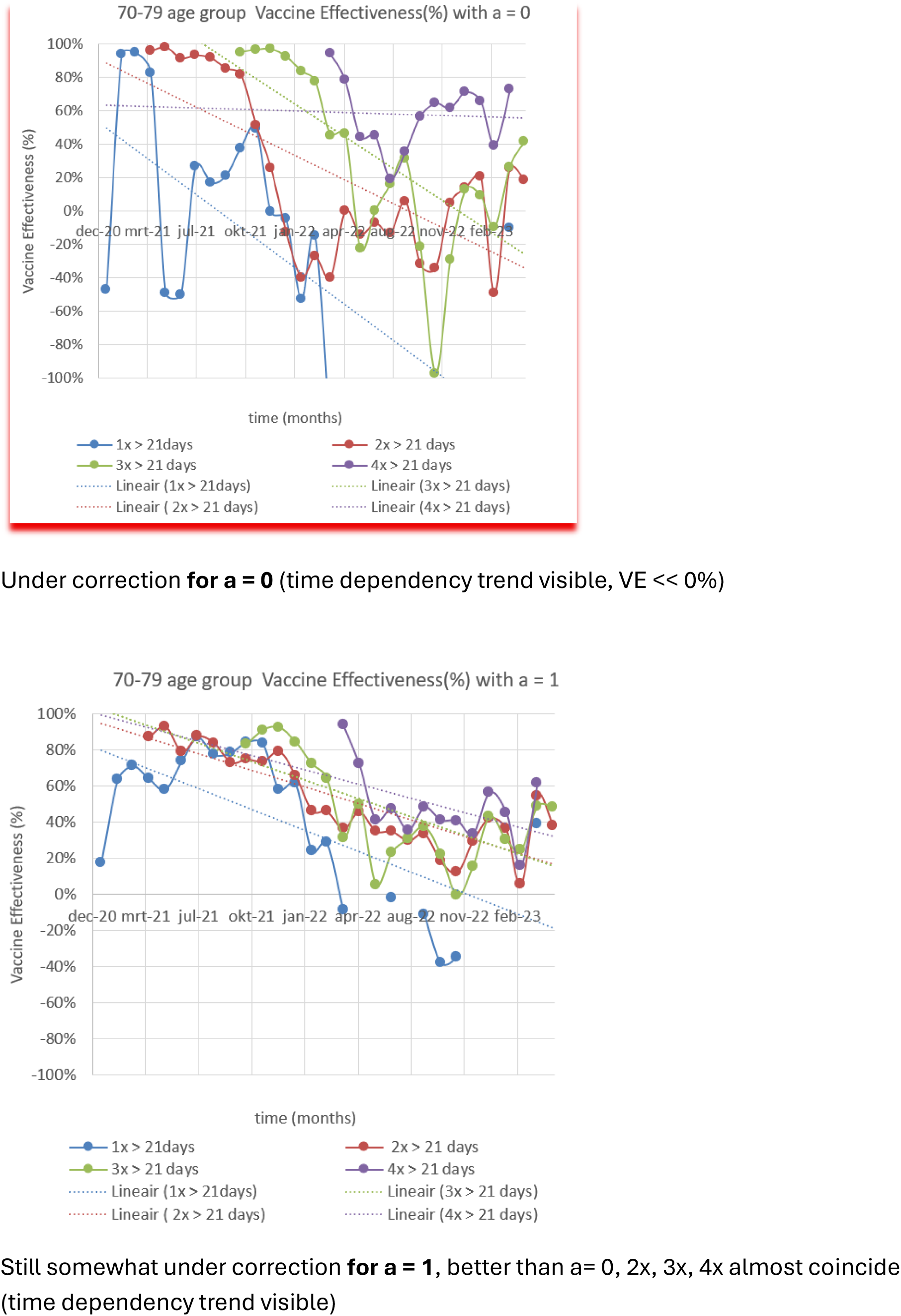

*Appendix V-C: Vaccine Effectiveness versus time: 70-79 years age group for different a-values (part 2*

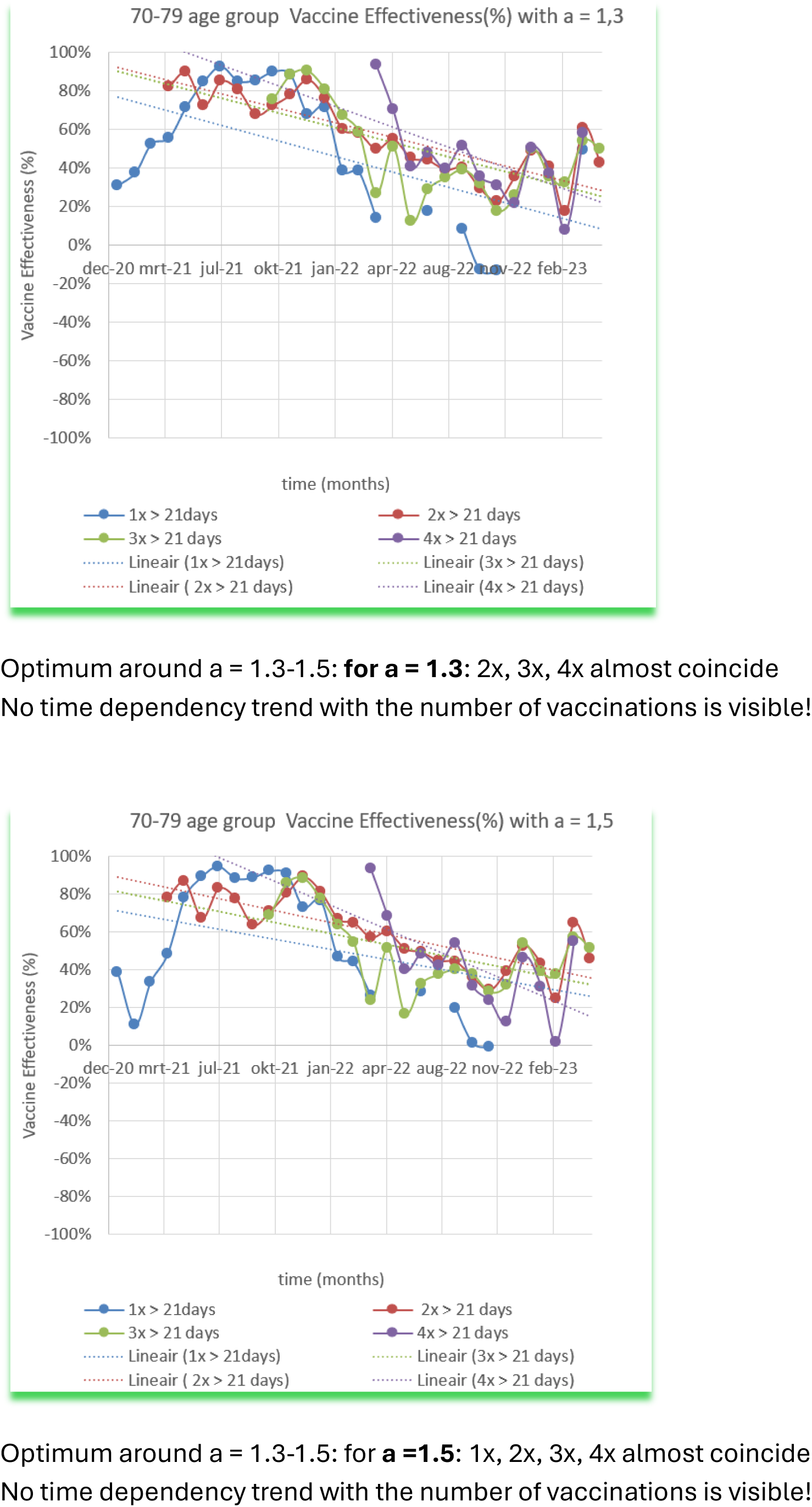

*Appendix V-C: Vaccine Effectiveness versus time: 70-79 years age group for different a-values (part 3)*

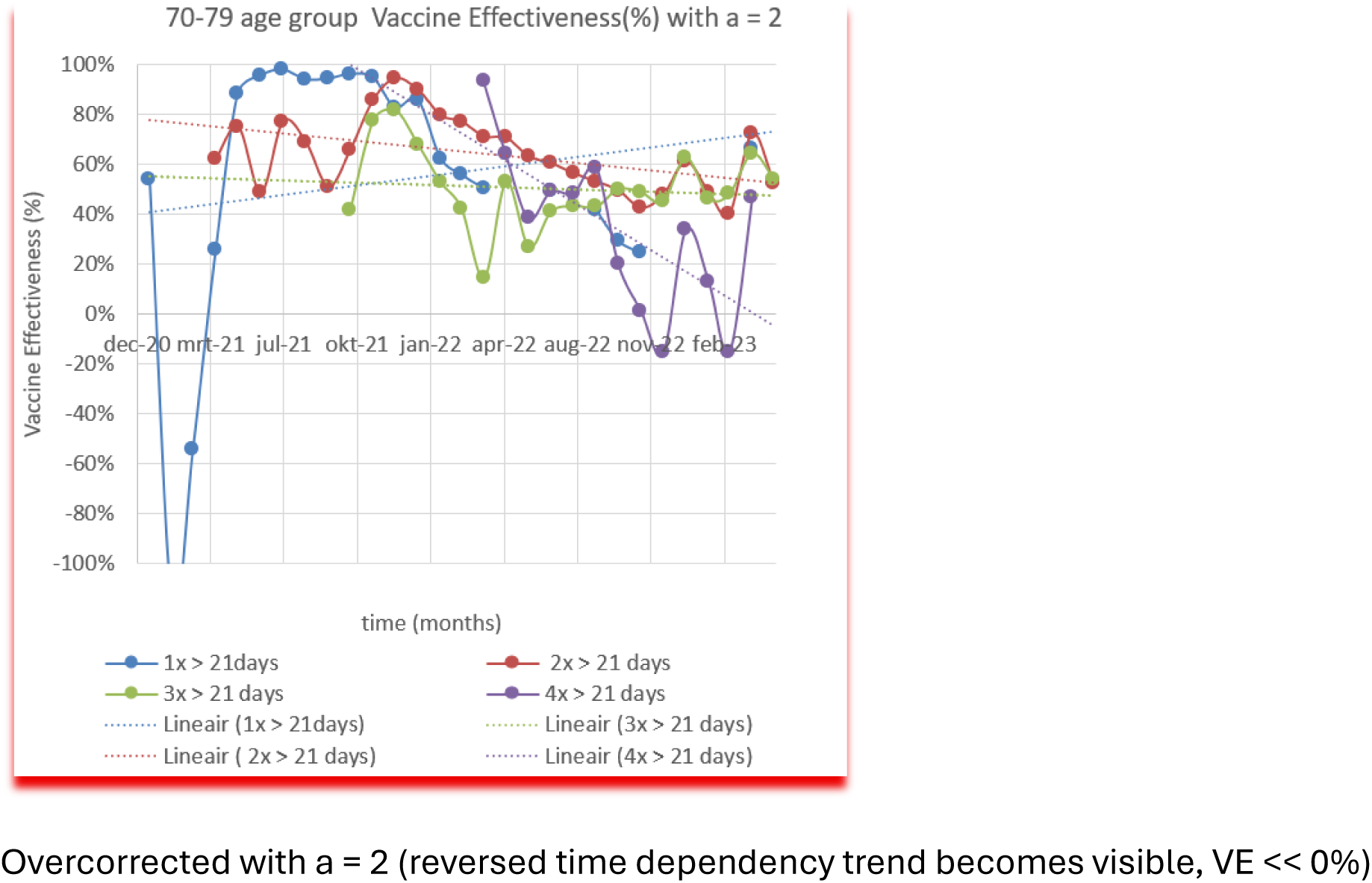

*Appendix VI-A: Vaccine Effectiveness for a = 0*, *1*, *1.5*, *2*, *2.5*, *3 in the 90+ age groups*

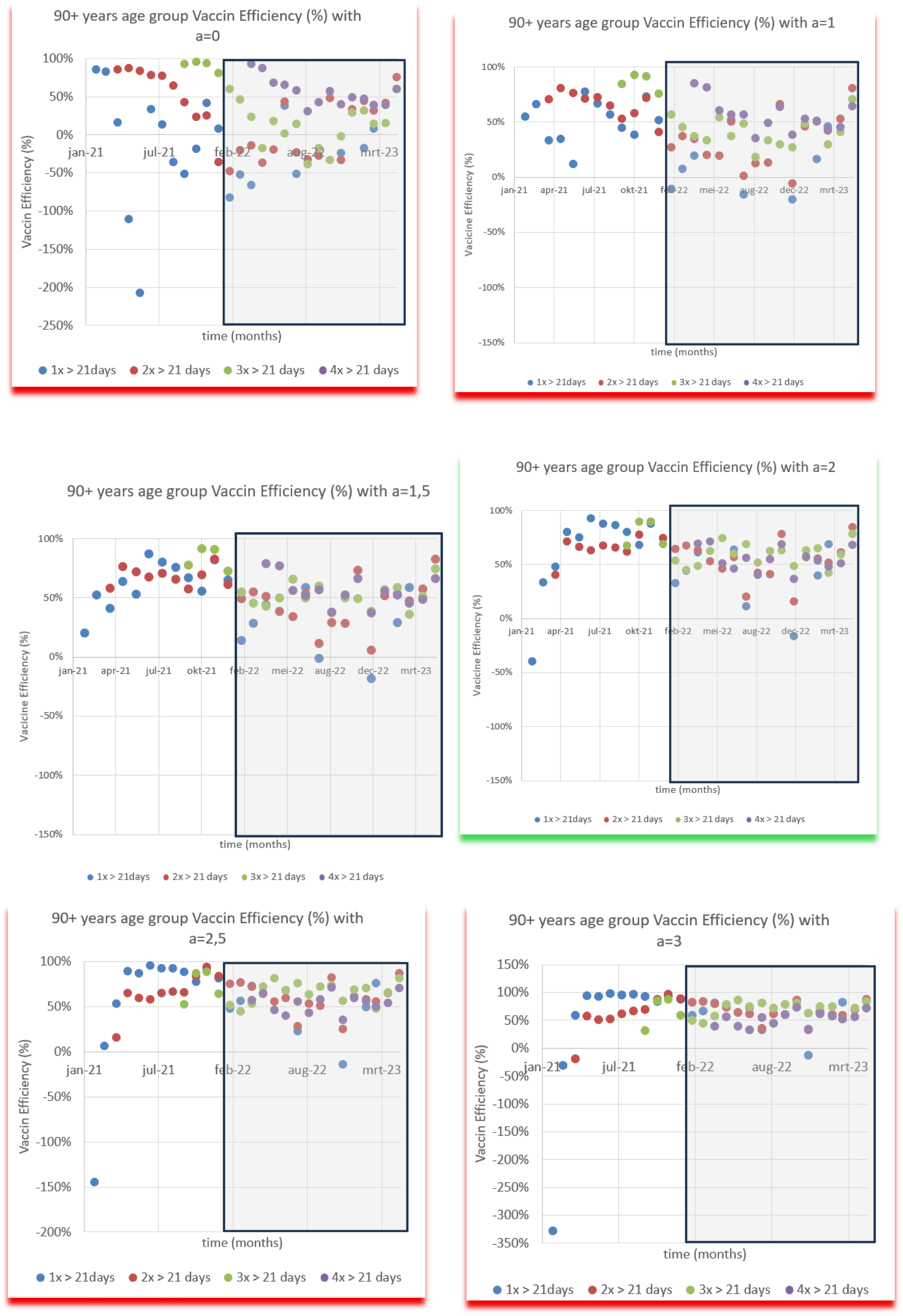

*Appendix VI-B: Vaccine Effectiveness for a = 0*, *1*, *1.5*, *2*, *2.5*, *3 in the 80-89 years age groups*

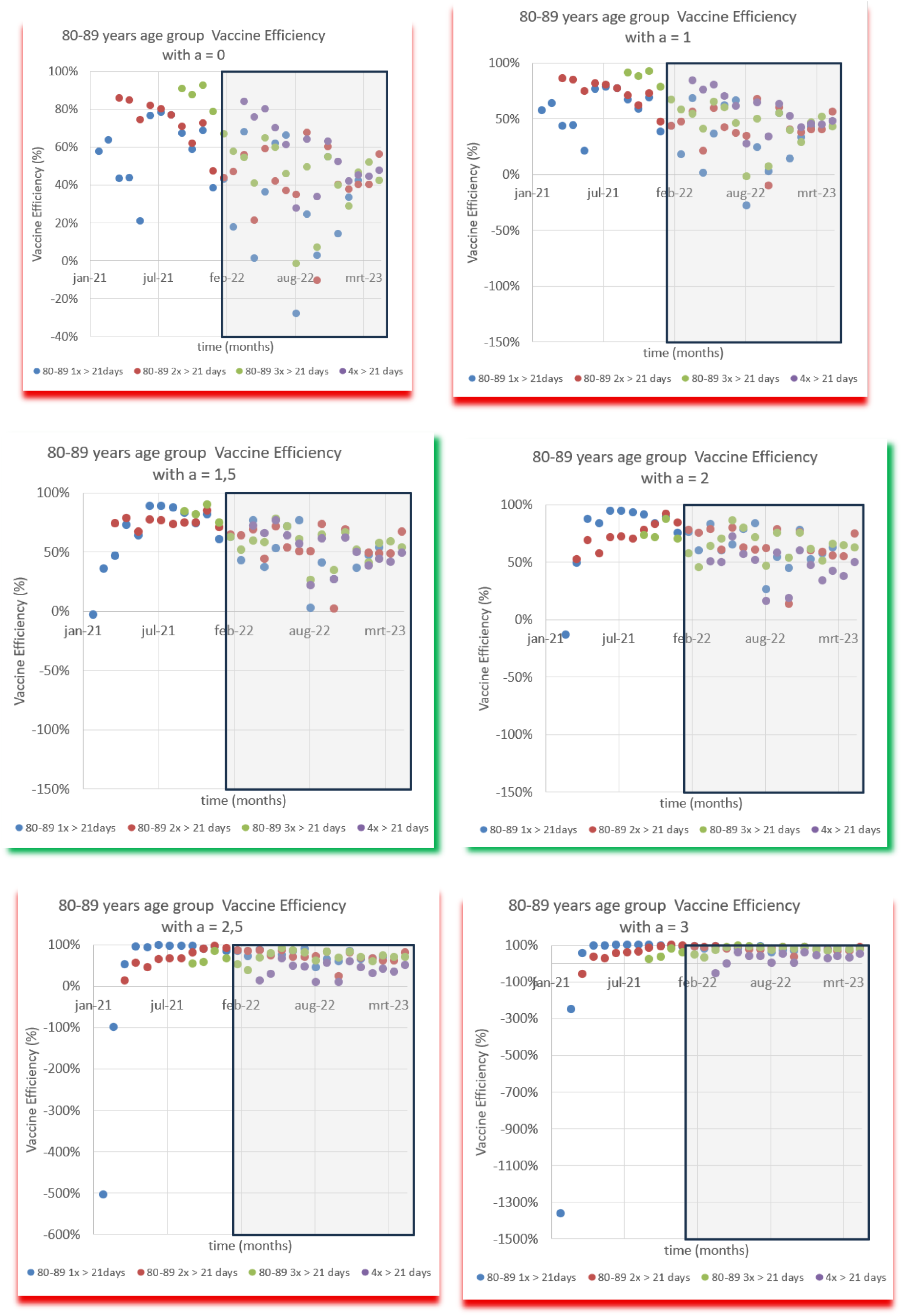

*Appendix VI-C: Vaccine Effectiveness for a = 0*, *1*, *1.5*, *2*, *2.5*, *3 in the 70-79 years age group*

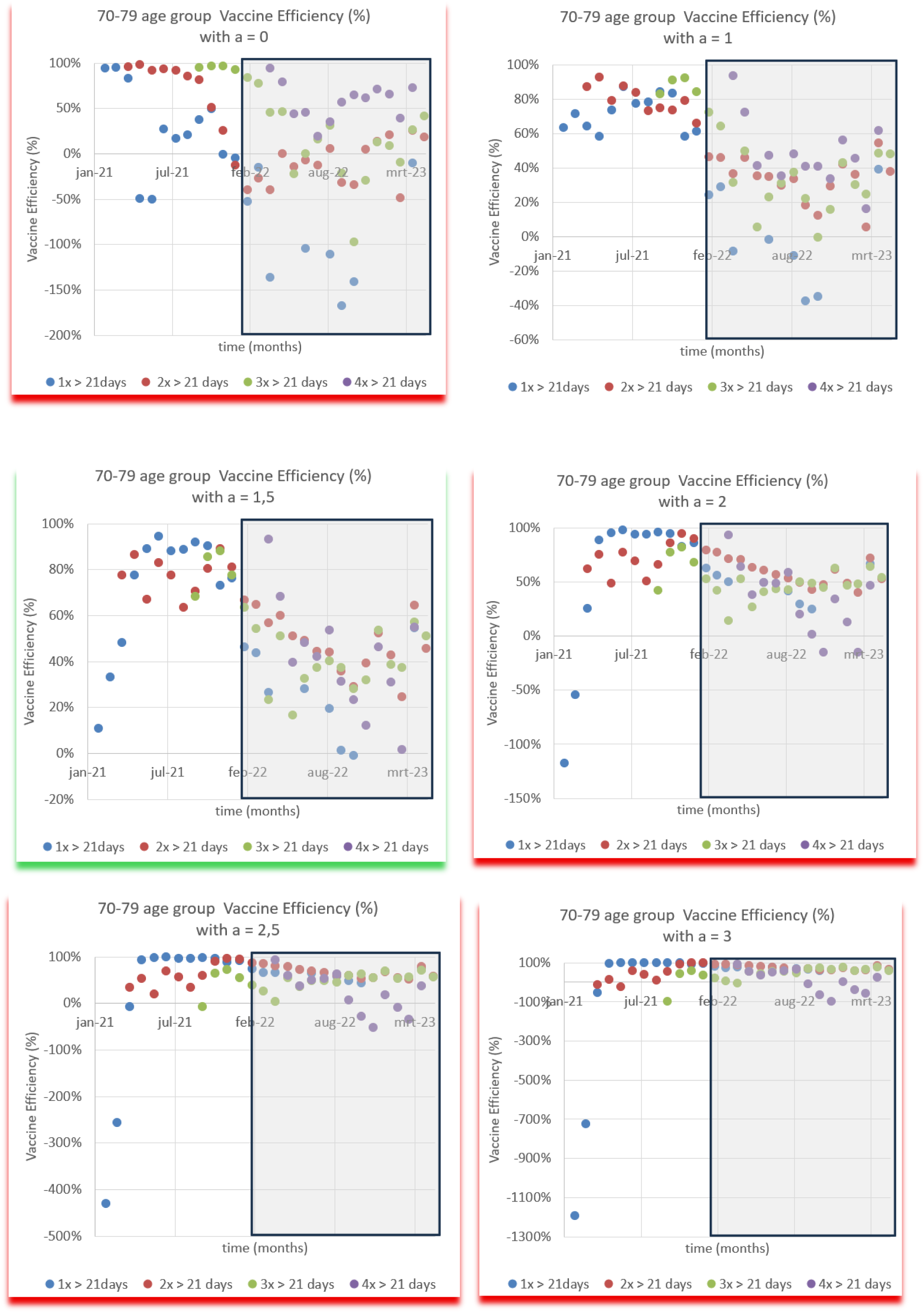

*Appendix VI-D: Vaccine Effectiveness for a = 0*, *1*, *1.5*, *2*, *2.5*, *3 in the 60-69 years age group*

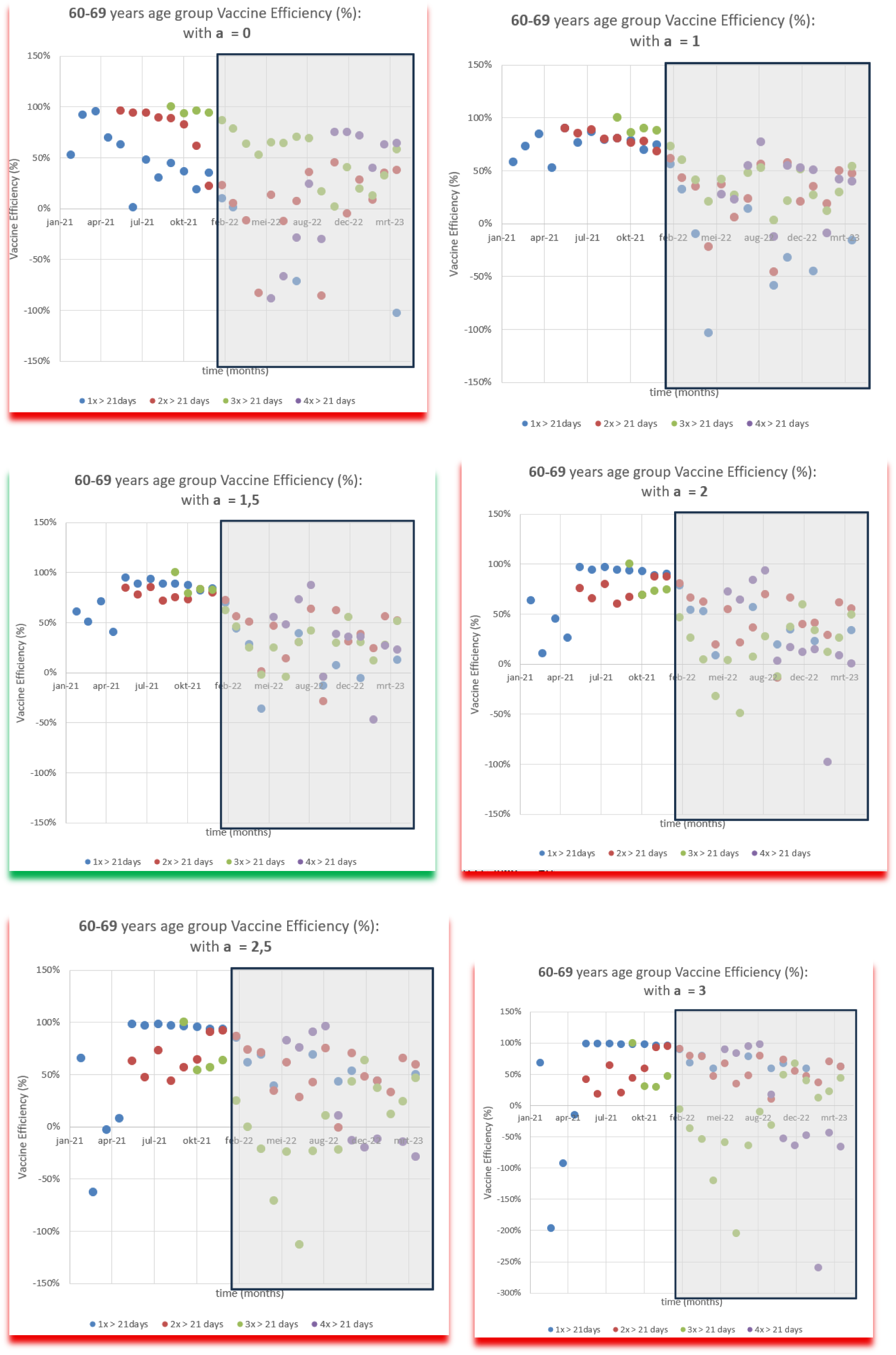

*Appendix VI-E: Vaccine Effectiveness for a = 0*, *1*, *1.5*, *2*, *2.5*, *3 in the 50-59 years age group*

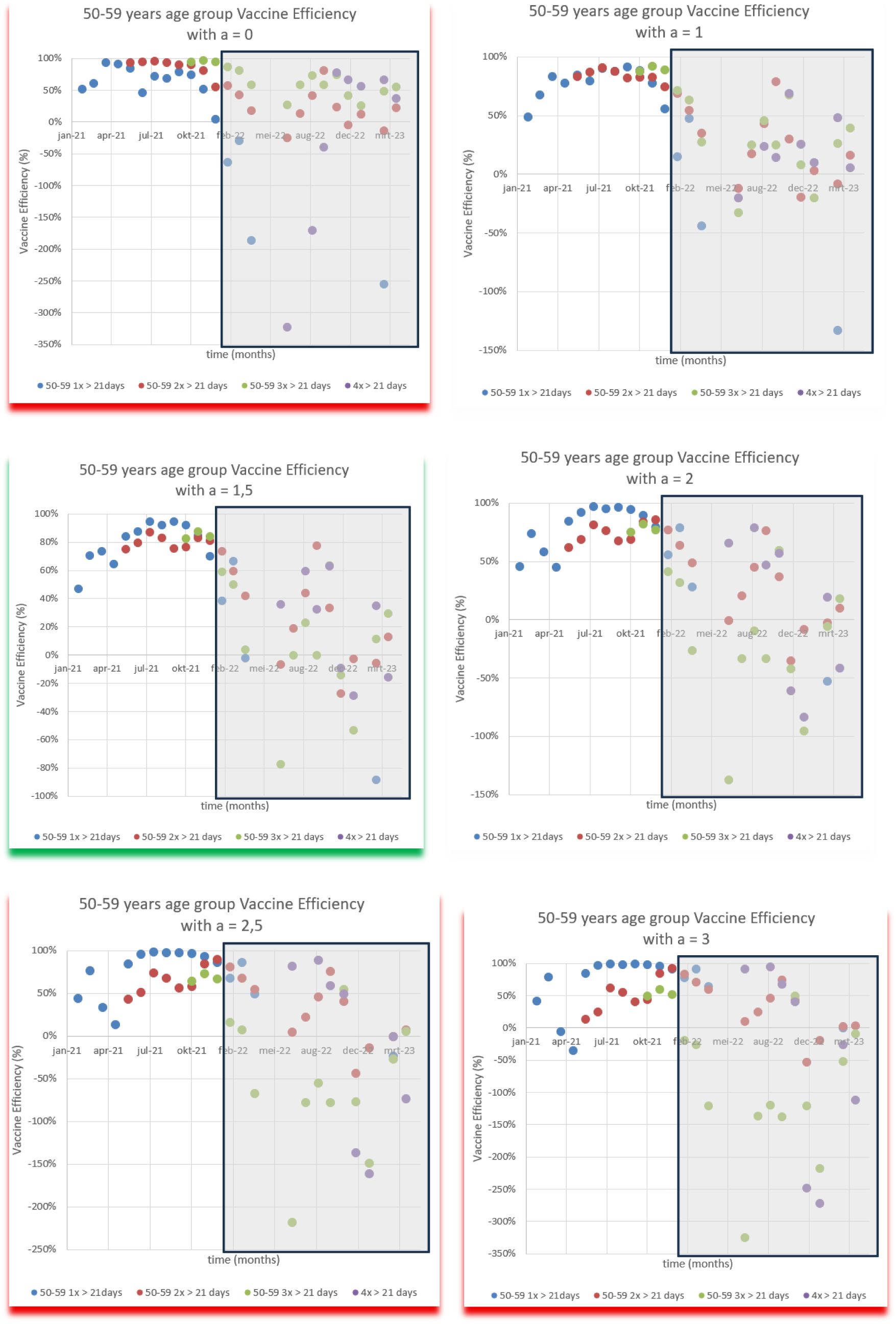

*Appendix VI-F: Vaccine Effectiveness for a=0,1,1.5*, *2*, *2.5 and 3 in the 40-49 years age group*

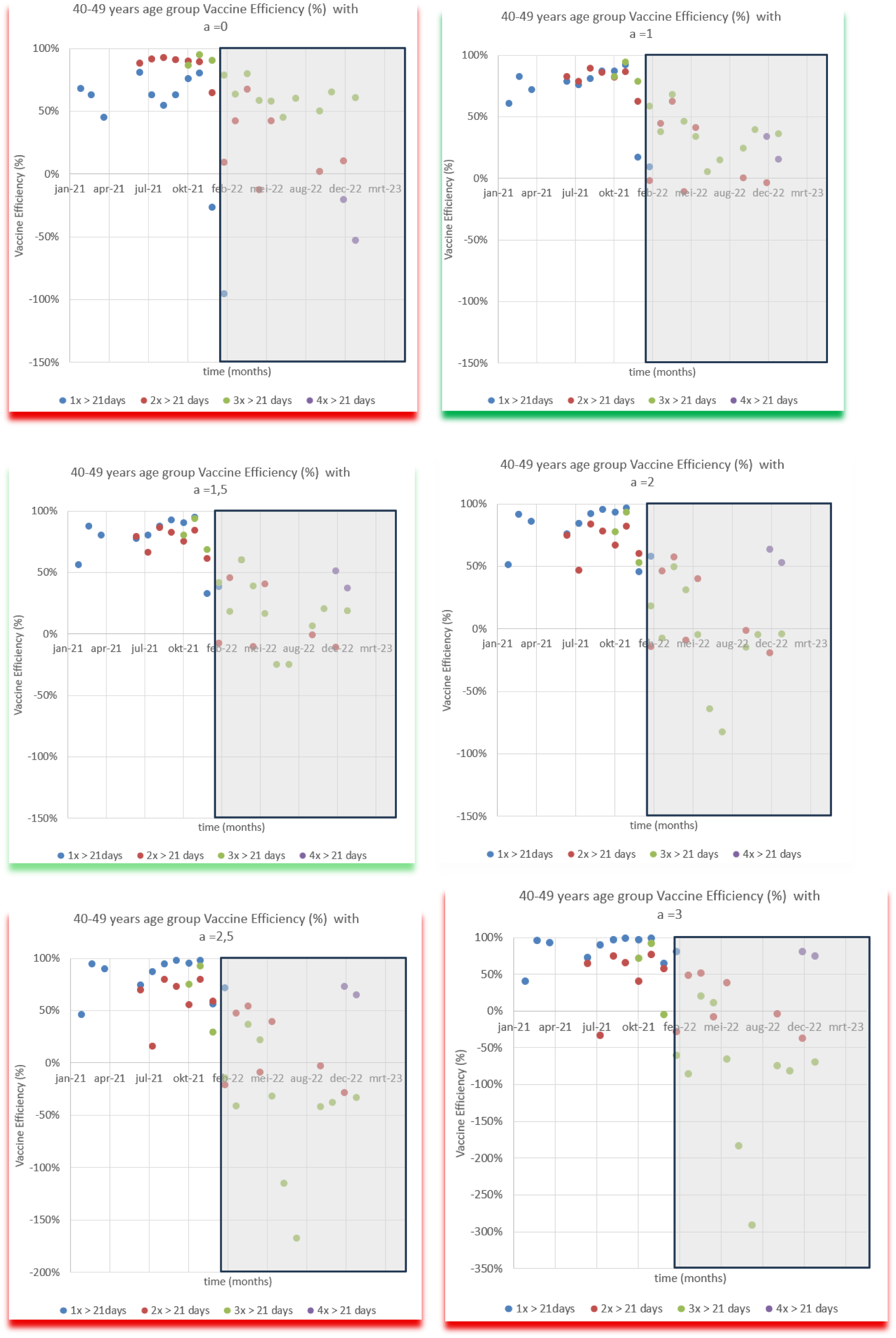

*Appendix VI-G: Vaccine EffectVeness for a=0,1,1.5,2,2.5,3a = 0*, *1*, *1.5*, *2*, *2.5*, *3a = 0*, *1*, *1.5*, *2*, *2.5 and 3 in the 18-39 years age group*

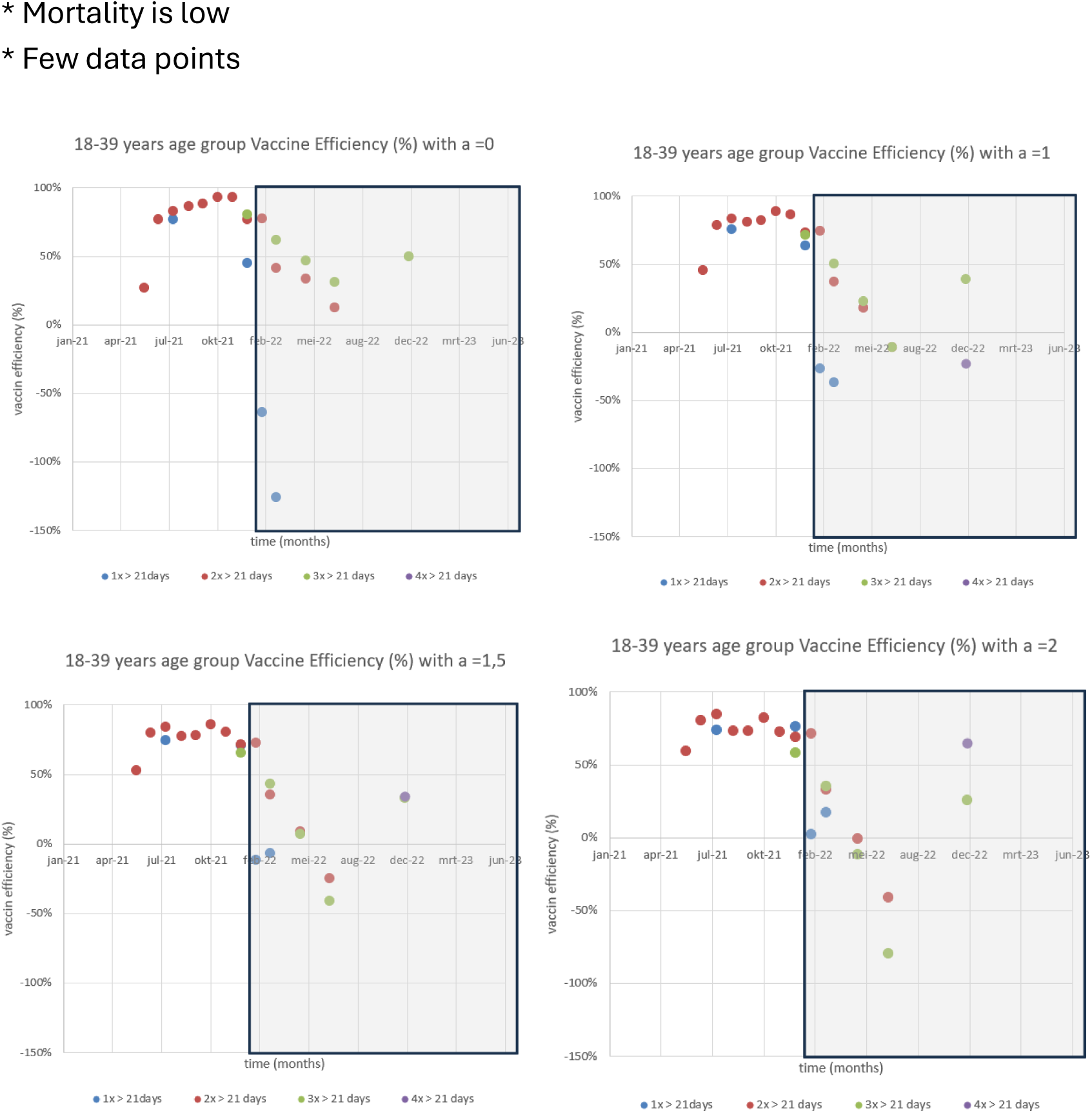

*VI-H: Analyses of Vaccine Effectiveness: Graphs for different a-values*

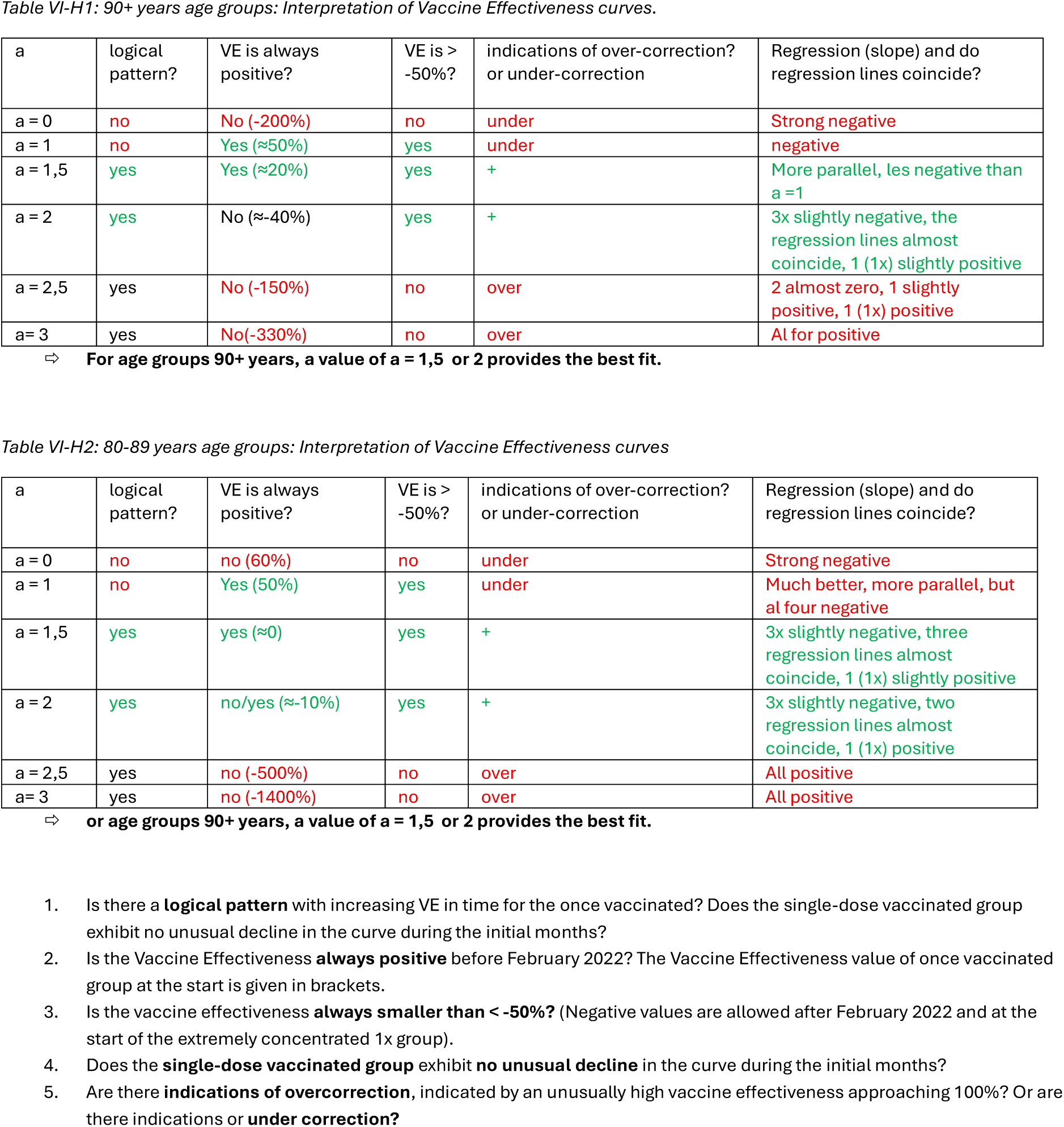

VI-H Analyses of Vaccine Effectiveness: Graphs for different a-values

**Table VI-H3:**
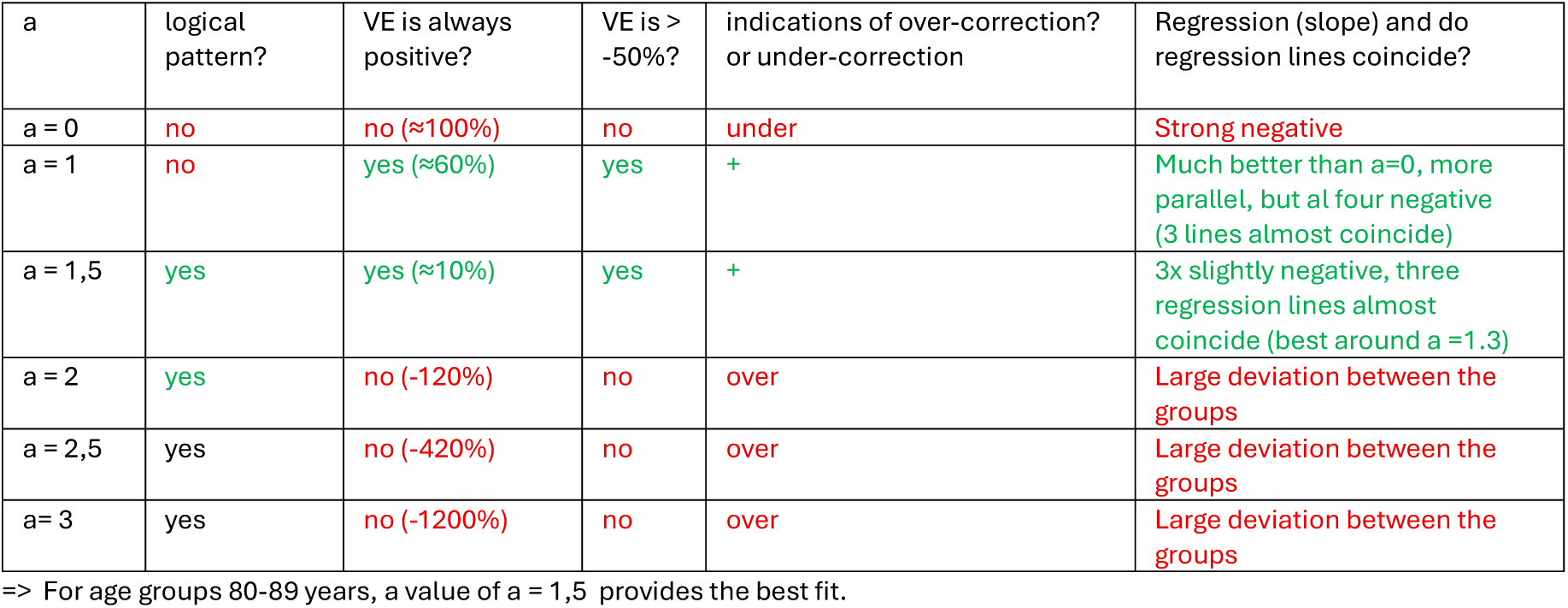
70-79 years age groups: Interpretation of Vaccine Effectiveness curves.

**Table VI-H4:**
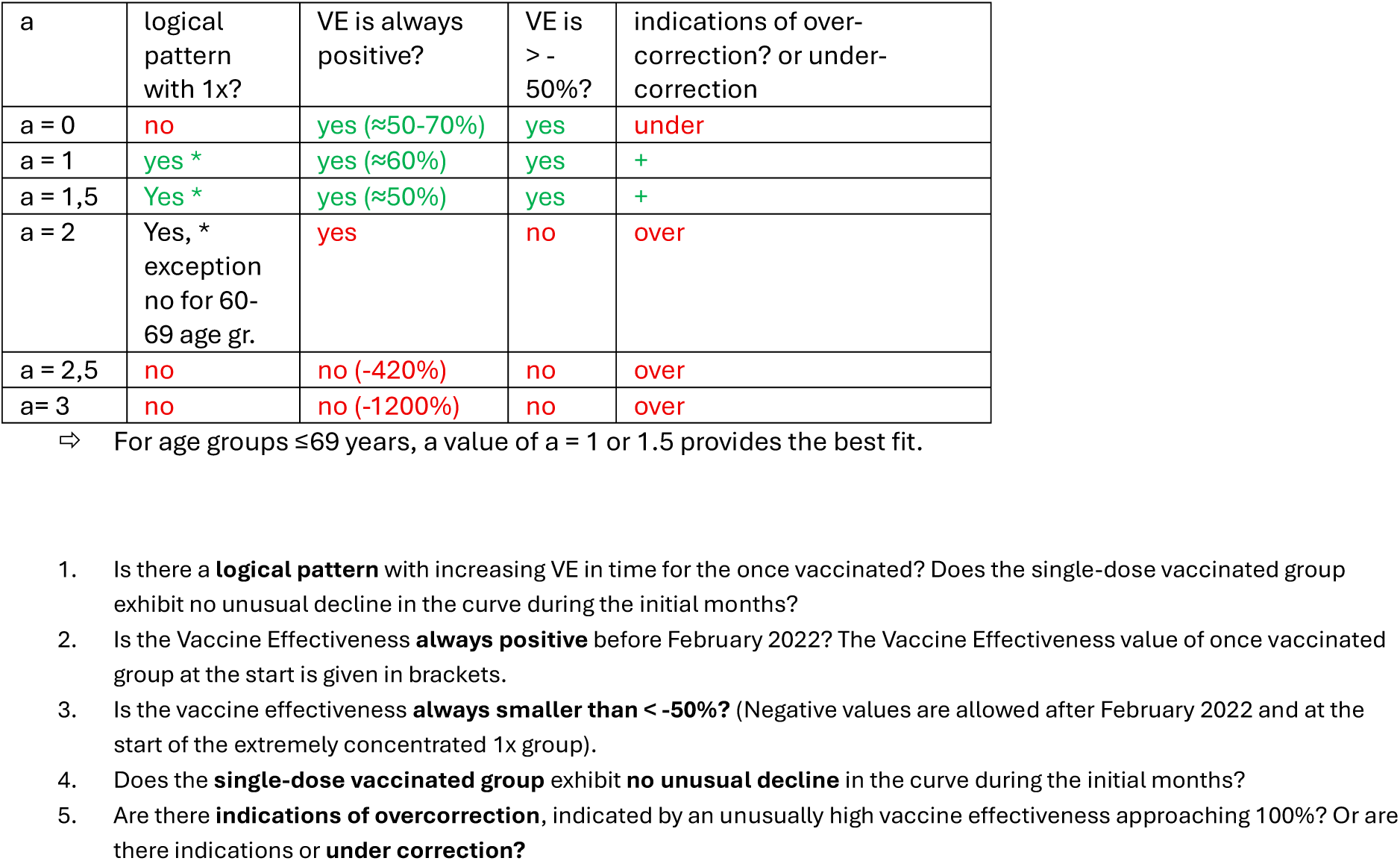
18-39, 40-49, 50-59 and 60-69 years age groups: Interpretation of Vaccine Effectiveness curves.

